# Modelling to infer the role of animals in *gambiense* human African trypanosomiasis transmission and elimination in DRC

**DOI:** 10.1101/2021.12.15.21267833

**Authors:** Ronald E Crump, Ching-I Huang, Simon E F Spencer, Paul E Brown, Chansy Shampa, Erick Mwamba Miaka, Kat S Rock

**Author notes:** These authors contributed equally to this work (Ronald E Crump, Ching-I Huang).

## Abstract

*Gambiense* human African trypanosomiasis (gHAT) has been targeted for elimination of transmission (EoT) to humans by 2030. Whilst this ambitious goal is rapidly approaching, there remain fundamental questions about the presence of non-human animal transmission cycles and their potential role in slowing progress towards, or even preventing, EoT. In this study we focus on the country with the most gHAT disease burden, the Democratic Republic of Congo (DRC), and use mathematical modelling to assess whether animals may contribute to transmission in specific regions, and if so, how their presence could impact the likelihood and timing of EoT.

By fitting two model variants – one with, and one without animal transmission – to the human case data from 2000–2016 we estimate model parameters for 158 endemic health zones of DRC. We evaluate the statistical support for each model variant in each health zone and infer the contribution of animals to overall transmission and how this could impact predicted time to EoT.

We conclude that there are 24/158 health zones where there is moderate or high statistical support for some animal transmission. However, – even in these regions – we estimate that animals would be extremely unlikely to maintain transmission on their own. Animal transmission could hamper progress towards EoT in some settings, with projections under continuing interventions indicating that the number of health zones expected to achieve EoT by 2030 reduces from 68 to 61 if animals are included in the model. With supplementary vector control (at a modest 60% tsetse reduction) added to medical screening and treatment interventions, the predicted number of health zones meeting the goal increases to 147/158 for the model including animals. This is due to the impact of vector reduction on transmission to and from all hosts.

**Author summary:** Elimination of African sleeping sickness by 2030 is an ambitious goal, not least because of the unclear role that animals might play in transmission. We use mathematical models, fitted to case data from DRC to assess and quantify the contribution of animals to the human case burden.

We found that 24/158 geographic regions included in this study had statistical evidence of animal transmission, although it appears extremely unlikely that animals could maintain transmission on their own. Animals could, however, delay elimination; using our model without animal transmission we predicted that 68 regions are expected to achieve elimination by 2030, whereas this reduces to 61 with animals. If vector control to reduce fly populations (which transmit the disease to and from hosts) are controlled in addition to medical interventions, then 147 regions are predicted to reach elimination by 2030 even with animal transmission.

## Introduction

Some infections which cause disease in humans can also be transmitted by non-human animal hosts, increasing transmission opportunities for the pathogen, and potentially hindering control of disease in humans or posing a threat to elimination or eradication. The lack of an animal reservoir is one of the numerous criteria listed as a requirement for a disease to be eradicable [1]. Guinea worm has become notorious for the surprises that can emerge as eradication is approached – despite the huge successes of reducing case reporting from 892,055 in 1989 to under 100 for the first time in 2015, case reduction subsequently stagnated [2] and the parasite has been identified in dogs [3] leading to speculation that this could impede eradication.

The mere presence of human infective parasites in animal populations is not sufficient to warrant immediate concern as this does not preclude the possibility that animals act as “dead-end” hosts, receiving infection themselves whilst not contributing to onward transmission. Even if onward transmission is possible, this alone does not constitute a reservoir; Haydon *et al*. [4] *provide a detailed description of what could be considered a maintenance* reservoir, specifying that such a population (or collection of populations) must be able to maintain infection on their own, even if infection were (temporarily) eliminated in humans. Of course, even some minor contribution from non-maintenance hosts could slow down progress to achieve an elimination or eradication goal, however control of infection in the human population should eventually enable complete removal of the infection from all human and non-human populations.

All is not lost for the Guinea worm programme [5], although undoubtedly the intensification in eradication efforts so close to zero have required a shift in how the programme must think about control, with uncertainty as to whether the goal can be met and a large price tag. Guinea worm is not alone in this uncertainty – another neglected tropical disease, the *gambiense* form of human African trypanosomiasis (gHAT), is an infection targeted for global elimination of transmission by 2030 [6], yet it remains unclear whether potential animal transmission could prevent this goal from being realised [7]. gHAT is a vector-borne infection, historically assumed to be solely transmitted to and from humans by blood-feeding tsetse. However, its close relative, *rhodesiense* HAT, is a known zoonosis with most infection occurring in animals [6].

There are a considerable number of reports of *Trypanosoma brucei gambiense* parasites in both wildlife and domestic livestock – Büscher *et al*. [7] *provide a concise recent summary and, in addition, a recent study reported T. b. gambiense* infections in domestic animals in Chad with 1.2–4.5% of those sampled testing positive in the three main foci [8]. Furthermore, experimental evidence of transmission of *T. b. gambiense* from animals back to humans, through the tsetse vector, has been documented [9], demonstrating that it is possible for animal-tsetse-human transmission cycles to exist. What these data are not telling us directly are whether this transmission pathway is occurring frequently, and to what extent it could be negatively impacting the concerted intervention efforts being made to control gHAT disease in humans.

Over recent decades, control methods for gHAT have primarily focussed on medical interventions in humans; either by diagnosis and treatment of symptomatic individuals at fixed health facilities (passive screening, PS), or through mass screening of at-risk villages (active screening, AS) [10]. There are also a range of methods available to target the tsetse vectors (traps, insecticidal targets, and ground or aerial spraying), although these have not been deployed at scale in many gHAT-endemic regions; in particular the Democratic Republic of Congo (DRC), which has the highest gHAT burden globally, currently only has large-scale tsetse control operations in a handful of its 189 gHAT-endemic health zones due to costs, other resource constraints and logistics over such a large geographical area [11]. Medical interventions, sometimes coupled with vector control, have resulted in a huge decline in transmission since 1998 with many countries achieving low or zero case reporting [2, 6]. The overall success to date certainly could lead us to have an optimistic outlook, yet the history of eradication programmes teaches us that some of the challenges we may face might not become readily apparent until we are very close to zero human cases.

The World Health Organization (WHO) have suggested the assessment of the role of animals as a key priority for gHAT modelling [10, 12]. Modelling studies conducted to date have been performed in a few distinct geographical locations, but have not tried to assess the questions surrounding the role of animal transmission across larger geographic regions.

In these studies, one used prevalence data from one focus in Cameroon in humans and various wild and domestic animal species at a single point in time, and found support for the presence of animal reservoirs but did not conclude with certainty whether gHAT could be maintained solely by transmission in animals [13]. Modelling studies in specific foci in the Democratic Republic of the Congo (DRC) and Chad fitted a gHAT model to longitudinal human case data, but not to animal data, and found similar statistical evidence for model variants including animal transmission compared to those with only human-tsetse transmission. Despite this inconclusive result the model fits were used to conclude that, if transmission from animals is occurring, the animals would not be able to maintain infection without humans, meaning they would not constitute a reservoir [14, 15]. A final study, focused on predictions for a focus in Guinea, did not try to quantify whether animal transmission was likely based on case data, but did conclude that, if there were some animal transmission, the likelihood of disease reemergence after stopping interventions would be high. This echoes related findings from a theoretical modelling study of gHAT which suggested that additional interventions, such as vector control, could be needed to curtail transmission if animals were able to acquire and transmit infection [16].

In the present study we seek to build upon this previous work to use data from longer time series and much larger geographic coverage to answer three primary questions: (1) What evidence is there for animal transmission based on longitudinal case data from across different health zones of DRC? (2) Are animals maintenance reservoirs? (3) If there *is* animal transmission, based on current trends, what will projections to 2030 and beyond look like? This third question in particular has important repercussions for planning policy and understanding the route to and sustainability of the 2030 EoT goal.

## Materials and methods

### Data

This study makes use of longitudinal human gHAT data for the DRC from the WHO HAT Atlas spanning the period 2000–2016 [17, 18]. The data contain information on the number of people screened each year in active screening campaigns by location, as well as those identified as cases through either active screening or passive surveillance.

Due to the required diagnostic and treatment algorithm during the data period, initially seropositive cases found in screening with the Card Agglutination Test for Trypanosomes (CATT) or a rapid diagnostic test (RDT) were subsequently confirmed using parasitological techniques before undergoing lumbar puncture to establish whether the infection had progressed to cross the blood-brain barrier. Finding either trypanosomes or elevated white blood cell (WBC) count (*>* 5 WBC/*µ*l) denotes a late stage or “stage 2” infection, meaning that patients had to be treated with nifurtimox-eflornithine combination therapy (NECT), as opposed to pentamidine which was used for patients in stage 1. In the WHO HAT Atlas data for DRC, staging information is typically not available prior to 2015, however this information is mostly present for 2015–16 and has previously proven to be informative when fitting mathematical models to gHAT data [19, 20]. Where available, staging information was included in the fitting, see Fig 1 and the Supplementary Information (S1 Materials and Methods).

**Fig 1.**
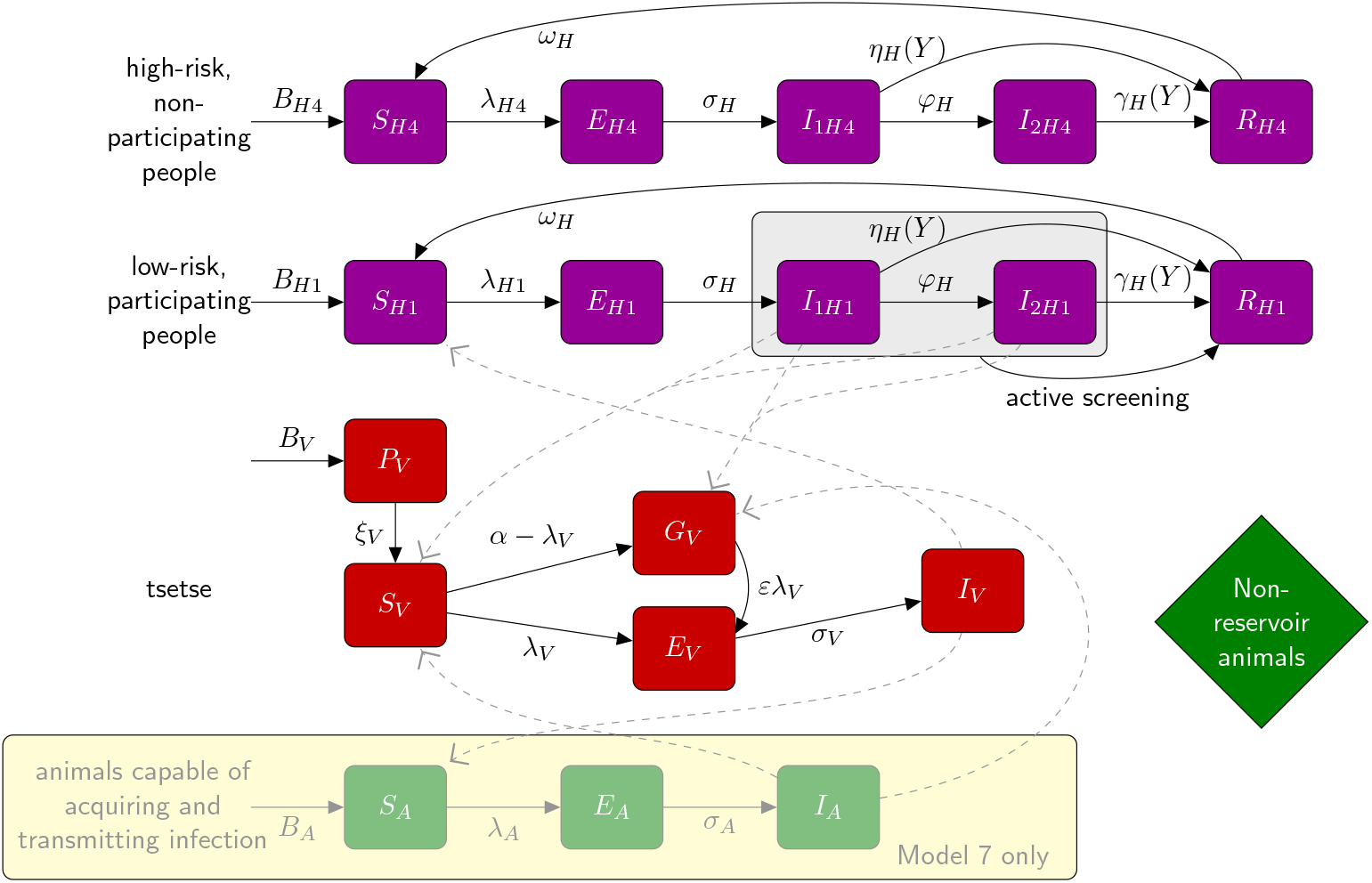
Warwick gHAT compartmental model variants 4 and 7. Purple boxes denote human infection states, with low-risk people who randomly participate in active screening denoted with subscript *H*1 and high-risk people who do no participate denoted with subscript *H*4 (we retain this notation to align with previously published version of this model). Tsetse are represented by red boxes and we explicitly include a pupal stage (*P*_*V*_) and differentiate between unfed (teneral) flies (*S*_*V*_) and non-infected but fed flies (*G*_*V*_) to incorporate reduced tsetse susceptibility following the first bloodmeal. For simplicity, the exposed category for tsetse is shown as a single compartment, *E*_*V*_ ; within the model this is subdivided into three compartments, *E*_1*V*_, *E*_2*V*_ and *E*_3*V*_, to model parasite development. The pathway relating to the animal infection and transmission, which is specific to Model 7, is highlighted with a pale yellow box. More details and model equations can be found in the Supplementary Information (S1 Materials and Methods).

For the purposes of this study we are interested in the health zone level aggregated data. Health zones are administrative units of approximately 175 000 people. We therefore take the same approach as utilised in our previous study to extract the data onto a recent shapefile for DRC [20] before aggregating by year, screening type (active or passive) and staging (stage 1, 2 or unknown) within health zone. Restricting to health zones containing at least 13 aggregated data points (either an active screening coverage of more than 20 people for the year or a non-zero passive detection), we find a total of 158 health zones with sufficient data for our analysis. This is 10 fewer health zones than included in Crump *et al*. (2021) where we only required 10 data points due to fitting a slightly less complex model.

Unfortunately there has been very limited sampling of animal populations for gHAT globally [7], and insufficient data are available for DRC to be able to fit our model to animal infection prevalence as well as human case reporting. Data collection for animal populations is challenging, especially if both wild and domestic animals are sampled. In addition to prevalence data, data on relative abundance of different animal species in relation to human or tsetse density, and feeding proportions of tsetse on animals are key pieces of information required to understand the full picture of potential tsetse-animal transmission cycles. Furthermore, at present there exists no 100% *gambiense* specific test for identifying conclusively that a non-human animal has *T. b. gambiense* infection rather than other animal trypanosomes, which are generally circulating at a much higher prevalence. Nevertheless there are modelling frameworks available that allow assessment of possible animal transmission without explicitly requiring animal data, which we will discuss below.

### Model variants

In the present study we present two previously developed models of gHAT, the first (denoted as “Model 4” for consistency with earlier work) is a solely anthroponotic transmission model incorporating transmission between the tsetse vector and human populations [20]. In Model 4, a proportion of bites are assumed to be taken on animals, however non-human animals are assumed to be dead-end hosts. This formulation effectively includes two possibilities (i) that animals do not acquire the parasite and are therefore not infected, or (ii) that animals can become infected, however they cannot transmit the parasite back to tsetse – both of these result in identical tsetse-human infection dynamics. In addition to a standard Ross-MacDonald formulation, there is also heterogeneity in the human population in terms of both risk of exposure to tsetse and participation in active screening. This model structure has been fitted to a number of different data sets [14,15,20,21], although certain aspects of the fitting procedure and simulation of passive surveillance have evolved since it was originally published.

The second variant we consider is “Model 7”. This model can be considered as an extension to Model 4, with the same risk and participation structure in humans. Model 7 also allows for a proportion of bites to be taken on animals which can both acquire infection from, and transmit infection back to, tsetse. This animal reservoir is in addition to the dead-end animal host population allowed for by Model 4. This formulation necessitates the addition of several extra model parameters: the effective animal reservoir density proportional to the human population, *k*_*A*_; the proportion of blood meals taken on these animals, *f*_*A*_; the mortality rate of an animal host, *µ*_*A*_, and the incubation rate of trypanosomes in animals, *σ*_*A*_.

Aside from the host and risk-structure mentioned above, numerous other details about the progression of disease in humans and interventions are encoded in the formulation of both model variants (see Supplementary Information S1 Materials and Methods for full model equations). In brief:

1. Passive detection rates can increase over time, we simulate this in all health zones of DRC in 1998 when the CATT became available for general use. In addition, we simulate subsequent improvements in provinces in which there is evidence of a pronounced increase in the proportion of passive cases found in stage 1 compared to stage 2; notably the former provinces of Bas Congo and Bandundu.
2. We assume that active screening occurs at the beginning of the year and that only low-risk people participate (as supported by previous modelling studies [14, 15]) although high- and low-risk people have equal passive detection rates.
3. We account for imperfect diagnostics in the active screening algorithm, with the standard screen-and-confirm process having a 91% sensitivity and very high but imperfect specificity (fitted independently in each health zone). In former Orientale province pre-2013, M’edecins Sans Frontières (MSF) were known to be operating an active screening algorithm based on CATT dilutions rather than parasitological confirmation and this is accounted for by reducing the comparative algorithm specificity for this period in this province. The use of video confirmation tools was introduced first in 2015 in Yasa Bonga and Mosango health zones in former Bandundu province and has subsequently been rolled out to the whole province. We simulate this as an increase to perfect specificity in 2018 for projections for other health zones in that province.
4. Whilst methods to control the tsetse vector are available, their wide-scale use was not implemented in DRC during the data period with the exception of Yasa Bonga health zone in former Bandundu province where Tiny Targets have been used as a tsetse intervention since 2015 [11]. We simulate bi-annual deployment of targets in this region, and furthermore we simulate future strategies with and without possible Tiny Target-based vector control (VC) in other health zones. (see Supplementary Information S1 Materials and Methods for equations relating to vector control).

### Fitting

Using a previously published Metropolis-Hastings Markov chain Monte Carlo (MCMC) algorithm, we fit each of Model 4 and Model 7 to the longitudinal human case data across the 158 health zones with sufficient gHAT data in DRC. Crump *et al*. [20] provide more details on the algorithm and presents the fitting for the human-only model (Model 4). Of the four extra parameters required for Model 7 we believe that *k*_*A*_, the effective animal density proportional to the human population, and *f*_*A*_, the proportion of blood meals taken on these animals, could be highly geographically heterogeneous and are therefore fitted within health zone. The other two Model 7-specific parameters, *µ*_*A*_, animal death rate, and *σ*_*A*_, animal incubation rate, are held constant (*µ*_*A*_ = 0.0014 days^−1^ and *σ*_*A*_ = 0.0833 days^−1^) across all health zones.

We use the same log-likelihood function as set out in Crump *et al*. [20] which matches model outputs of human case reporting to the longitudinal data for both active and passive detection modes, including staged reporting if this is available in the data. The full equations are found in the Supplementary Information (S1 Materials and Methods) and include overdispersion in case detection to account for larger variance than expected under the binomial distribution. This log-likelihood function remains unchanged as no data on animal prevalence are available in the present study, however the two additional fitted parameters in Model 7 are accounted for through their impact on the expected human case numbers.

#### Priors

Priors for all fitted parameters common to both models are given in the Supplementary Information (S1 Materials and Methods). For the effective animal density, *k*_*A*_, a Γ(1.26, 19.3) prior distribution was used; with a mode of 5.0, and 2.5% and 97.5% quantiles equal to 1.2 and 82.4, respectively. The proportion of blood meals taken on these animals, 0 *≤ f*_*A*_ ≤ 1 − *f*_*H*_, where *f*_*H*_ is the proportion of blood meals taken by tsetse on humans, had a flat/uniform prior over this range as any value seemed biologically plausible, and would be very dependent on tsetse feeding preference and animal abundance.

### Model comparison

Following the fitting of each model, model comparison was performed using Bayes factors (*K*_*ij*_), which are ratios of the model evidence (or marginal likelihoods) under models *i* and *j*. For example, if for a particular health zone, *K*_47_ = 1.2, then Model 4 is slightly favoured over Model 7, but the evidence is weak. Conversely if *K*_74_ = 102 then Model 7 is decisively favoured over Model 4. Categorisations of Bayes factor value interpretations are given alongside results.

Importance sampled estimates of the model evidence for each model in each health zone were obtained following the methodology of Touloupou *et al*. [22], utilising a defense mixture [23]: in our study a weighted combination of a multivariate Gaussian mixture fitted to the 2 000 samples from the joint posterior distribution of the fitted model parameters (weight = 0.95), and the prior distributions of the fitted model parameters (weight = 0.05). See the Supplementary Information (S1 Materials and Methods) for additional details.

### Host-specific reproduction numbers

For Model 7 we can also assess the relative contribution of non-human animal reservoirs to transmission, and assess their potential competence to maintain infection in the absence of transmission in humans. To do this, host specific, i.e. human or animal, reproduction numbers were calculated using a next generation matrix approach [13] with a design matrix identifying contributions from the two host pathways. See the Supplementary Information (S1 Materials and Methods) for full details.

### Projections

The results of the model fitting for Models 4 and 7, in the form of 1 000 realisations from the joint posterior distribution of all fitted parameters, were used to make projections under two main strategies. These are a subset of those reported in the paper on projections for Model 4 [24]. In both strategies, active screening was continued beyond 2016 at the mean level observed in that health zone in the period 2012 to 2016, this was chosen as reflecting an achievable level of active screening in that locale with recent, if not current, resource availability. We present active screening at higher coverage (the historical maximum in each health zone during 2000–2016) as a sensitivity analysis in our graphic user interface (GUI, online https://hatmepp.warwick.ac.uk/animalfitting/v1/), but do not focus on these results in the main manuscript. Passive screening was assumed to continue at the 2016 level of effectiveness. The two strategies differ with regards to vector control (VC): in the basic strategy no VC was carried out, while in the second VC was assumed to be implemented from 2020, with annual tsetse population reductions of 60%, which is a low, conservative estimate of potential VC effectiveness when this strategy is deployed at scale; this value is below tsetse reductions reported elsewhere in programme or study implementation which have reached up to 99% reduction in specific settings [11,15,25,26]. Whilst we use this lower bound for VC effectiveness in the main manuscript, 80% and 90% tsetse reductions are presented as sensitivity analysis in our GUI to show other reductions could impact transmission and reporting further. The VC exception is for Yasa Bonga health zone in the former Bandundu province, where VC was actually implemented from 2015. Note that even in Yasa Bonga where 90% tsetse reductions were reported [11], we assumed the reported 90% tsetse reduction after the first year.

The Matlab code used to perform the MCMC fitting to historical data, calculate host-specific reproduction numbers, approximate the model evidence and make projections is available from https://osf.io/3xadf/.

## Results

### Fitting and Bayes factors

We fitted both model variants to the health zone level data for 158 health zones. Fig 2 show results for two example health zones, Bokoro in former Bandundu province, and Tandala in former Equateur province, and results for all other health zones can be viewed online in our companion GUI (https://hatmepp.warwick.ac.uk/animalfitting/v1/). Quantiles for reported active and passive case numbers were of 10 000 stochastic samples, 10 samples for each of 1 000 random samples from the joint posterior distribution of the fitted model parameters. The estimated number of new human infections are taken from the solutions of the ordinary differential equations for the model for each of the 1 000 posterior parameter sets.

**Fig 2.**
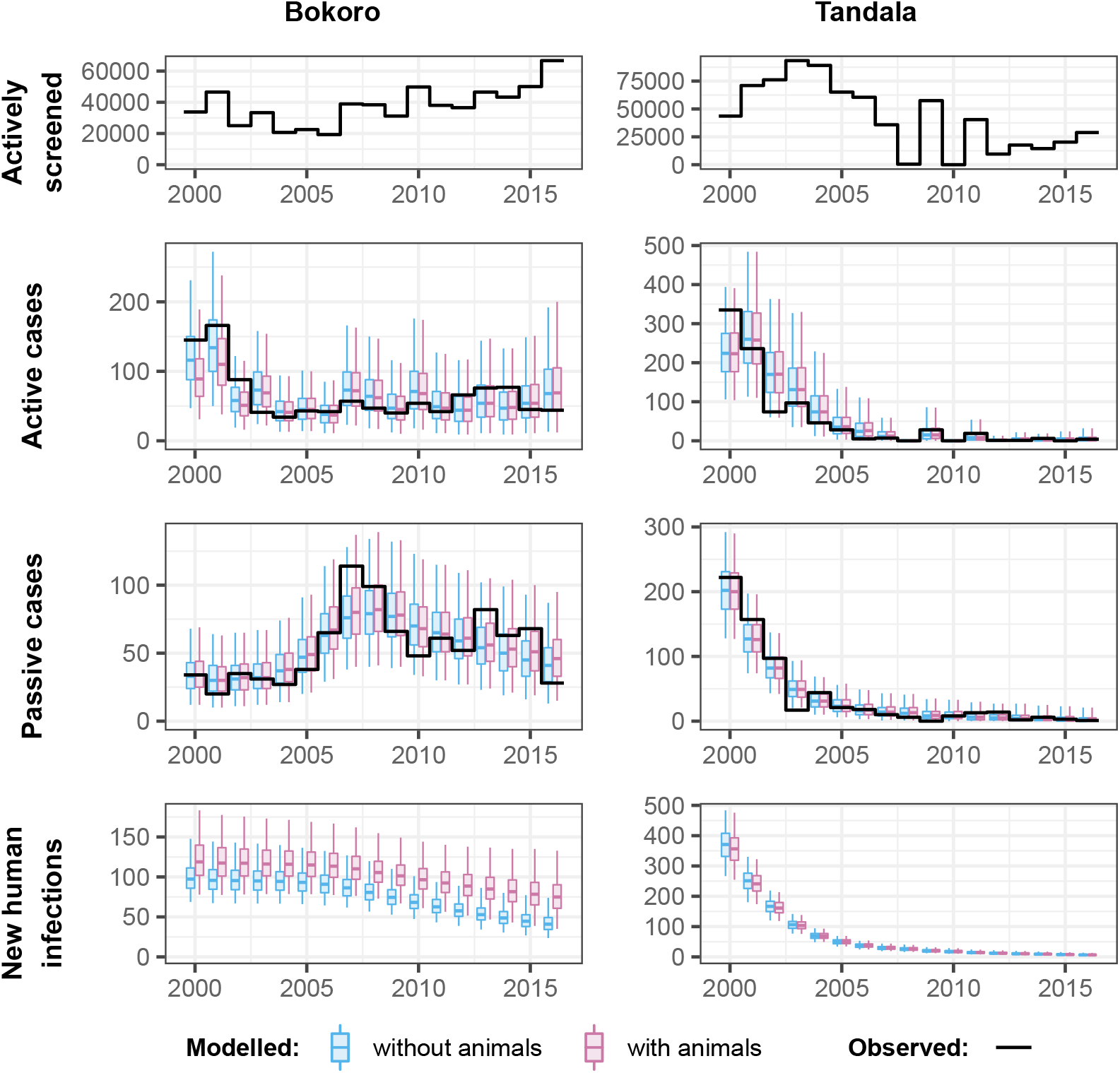
Fits to historical case data for Bokoro and Tandala health zones, for models with (in pink) and without (in blue) animals contributing to gHAT transmission. Reported data are shown as a solid black line and each boxplot shows the 2.5% and 97.5% quantiles of the model fit as the extreme point of the whiskers, 25% and 75% quantiles delimiting the box, and the median as the mid-line of the box.

The fit to both the active and passive case numbers is shown to capture the reported case trends well, including the observed ‘humped’ shape to the passive detection in Bokoro, which is also notable across other health zones of the former Bandundu province; this shape is associated with improvement to passive detection rates from both stage 1 and 2 of the disease. Both Model 4 and Model 7 fit very similarly to the observed data points. However in Bokoro health zone, which has strong support for the model with animal transmission, there are more new human infections each year under Model 7, and also more unreported deaths (deaths not shown in the figure).

To compute the support for each of the model variant following fitting we categorised the Bayes factors in a widely accepted way: Weak (‘Barely worth mentioning’), 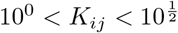 Substantial, 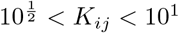 Strong, 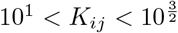 Very Strong, 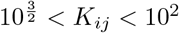 and Decisive, *K*_*ij*_ > 10 (*K*_74_ or *K*_47_ [27]. If *K*_74_ > 1, the categorisation for *K*_74_, indicating support for the model with animal transmission was displayed, otherwise the categorisation for *K*_47_ was used. A map of our results is shown in Fig 3. The majority of health zones in which the model with animal transmission had decisive support (24 health zones in which 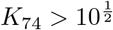) are in the former provinces of Equateur and Kasai Oriental (12 and 8 health zones, respectively). Many health zones had weak support for the model without animals.

**Fig 3.**
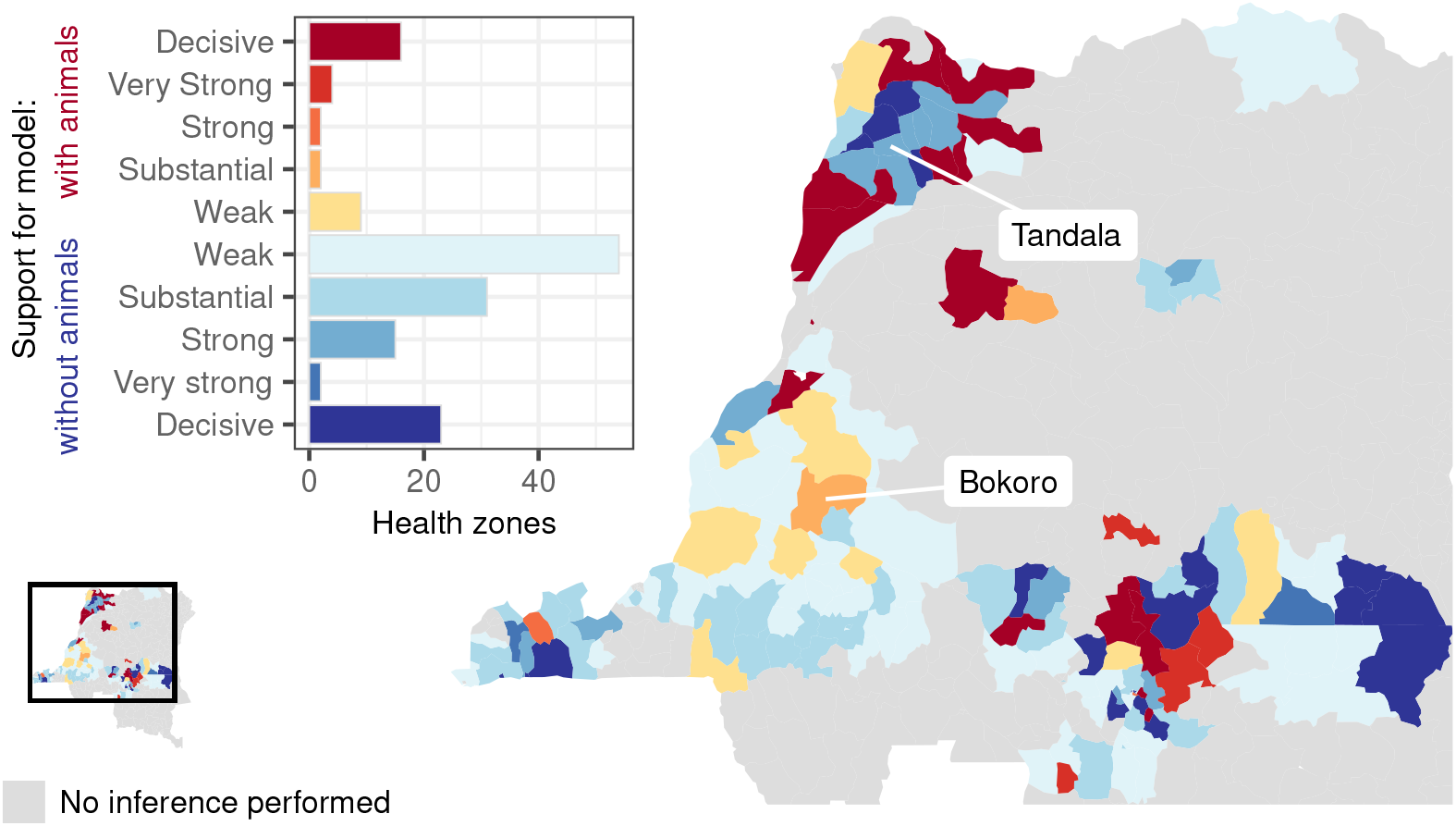
Support for model either with or without animals contributing to transmission of gHAT. Levels of support taken from whichever of the two Bayes Factors (that is, with either evidence for the model with or without animal transmission as the denominator) exceeded 1. Weak (‘Barely worth mentioning’), 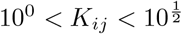 Substantial, 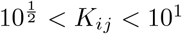 Strong, 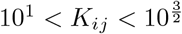 Very Strong, 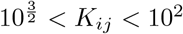 and Decisive,*K*_ij_ > 10^2^ (*K*_74_ or *K*_47_ if the model with or without animal transmission is favoured, respectively). Health zones used as a examples in later figures, Bokoro and Tandala, are indicated. Shapefiles used to produce these maps are available under an ODC-ODbL licence at https://data.humdata.org/dataset/drc-health-data.

Next, we used the posteriors from fitting Model 7 to assess the host-specific reproduction numbers. Fig 4 shows the 2 000 estimates of the reproduction rate in humans 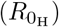 and animals 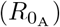 against one another for our two example health zones: Bokoro and Tandala. These posterior parameter samples can be summarised to give the probability that transmission in the animals only (where 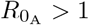 and 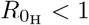), in humans only (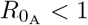 and 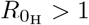), in either humans or animals (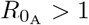 and 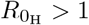), or in humans and animals (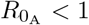 and 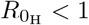) would be capable of maintaining transmission of gHAT. There are no parameter sets in Bokoro or Tandala with a non-zero probability of animals being a maintenance host. In these health zones there is non-zero probability that humans could be a maintenance host for gHAT (0.03 in Bokoro, 0.70 in Tandala) or that transmission in both humans and animals is required to maintain gHAT transmission (0.97 in Bokoro, 0.3 in Tandala). These host-specific reproduction number results align with the substantial support for the model with animal transmission in Bokoro and strong support for the model without animal transmission in Tandala (see Fig 3).

**Fig 4.**
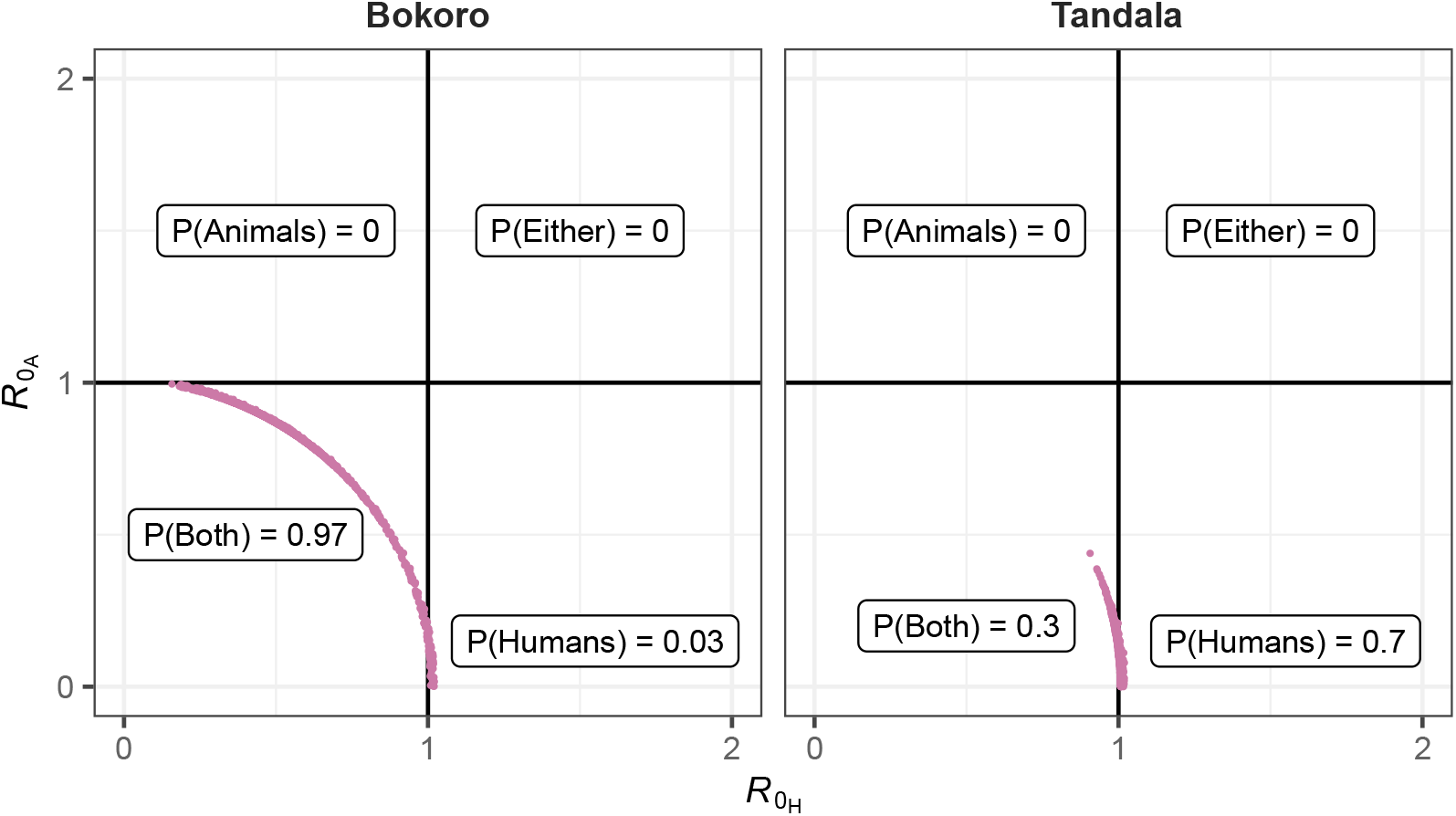
Basic reproduction number for animals 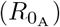 and humans 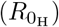 for two example health zones, Bokoro and Tandala, calculated for each of 2 000 posterior parameter sets of the model including animal transmission. The probability that transmission in animals only 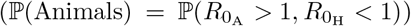, humans only 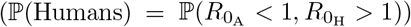, both animals and humans 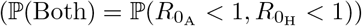, or either animal or humans 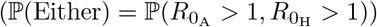 are sufficient to maintain on-going gHAT transmission are indicated.

We can do the same calculation for all health zones across DRC. In 132 of the 158 health zones the most probable scenario was that transmission in both humans and animals would be required to maintain gHAT transmission. Fig 5 shows the results of the host-specific basic reproduction number calculations exemplified in Fig 4 summarised for all 158 analysed health zones. There were nine health zones in which some posterior samples resulted in 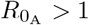, and hence non-zero probabilities that transmission in animals alone would be sufficient to maintain gHAT transmission. The highest value of this probability, 0.045, was observed in Lubunga 2 health zone in the former Kasai Occidental province. For Lubunga 2 health zone; the probability that both human and animal transmission are required for maintenance was 0.892, the probability that humans are a maintenance host was 0.063, and furthermore there was weak support for the model with an animal reservoir.

**Fig 5.**
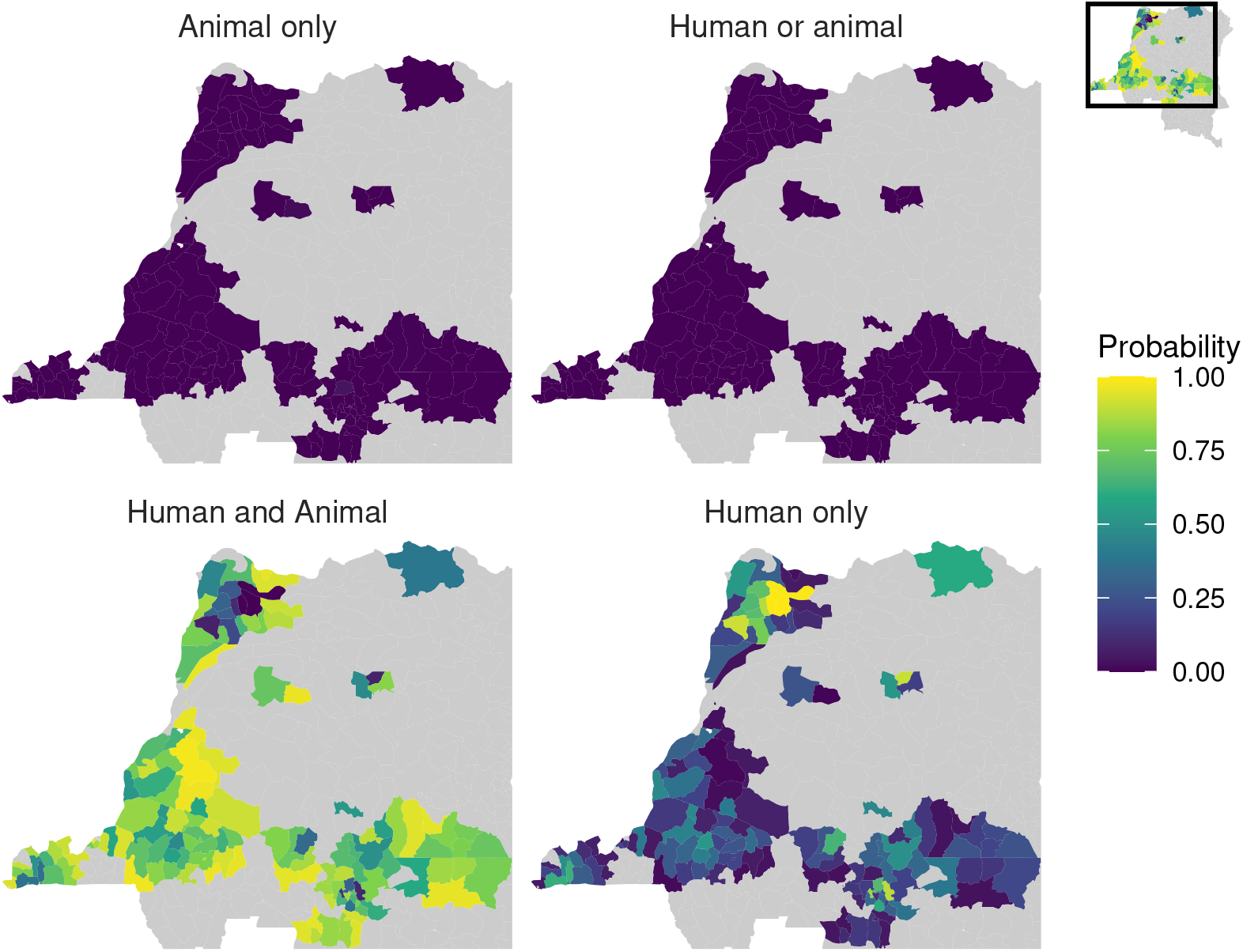
Each subplot shows the probability the maintenance reservoir in each health zone of DRC consists of: just animals, either humans or animals, humans and animals combined or just humans. In the bottom two subplots, humans are required to have sustained infection and animals could not maintain transmission of gHAT in the absence of human infection. Shapefiles used to produce these maps are available under an ODC-ODbL licence at https://data.humdata.org/dataset/drc-health-data.

### Projections using models with and without animal transmission

We next used the posterior parameterisation of each health zone to predict future trends in case reporting and transmission. Bokoro and Tandala had averages of 24% and 7% annual active screening coverage from 2012–2016, respectively. Fig 6 shows the projected impact of continuing active screening alongside passive screening at the current level of effectiveness at this level from 2017 to 2040, on reported cases and human infections. With or without the presence of animal transmission, there are downward trends in these measures in these health zones. However there are more new active and passive cases reported each year, and more new human infections each year, under Model 7, with an impact on the achievement of EoT.

**Fig 6.**
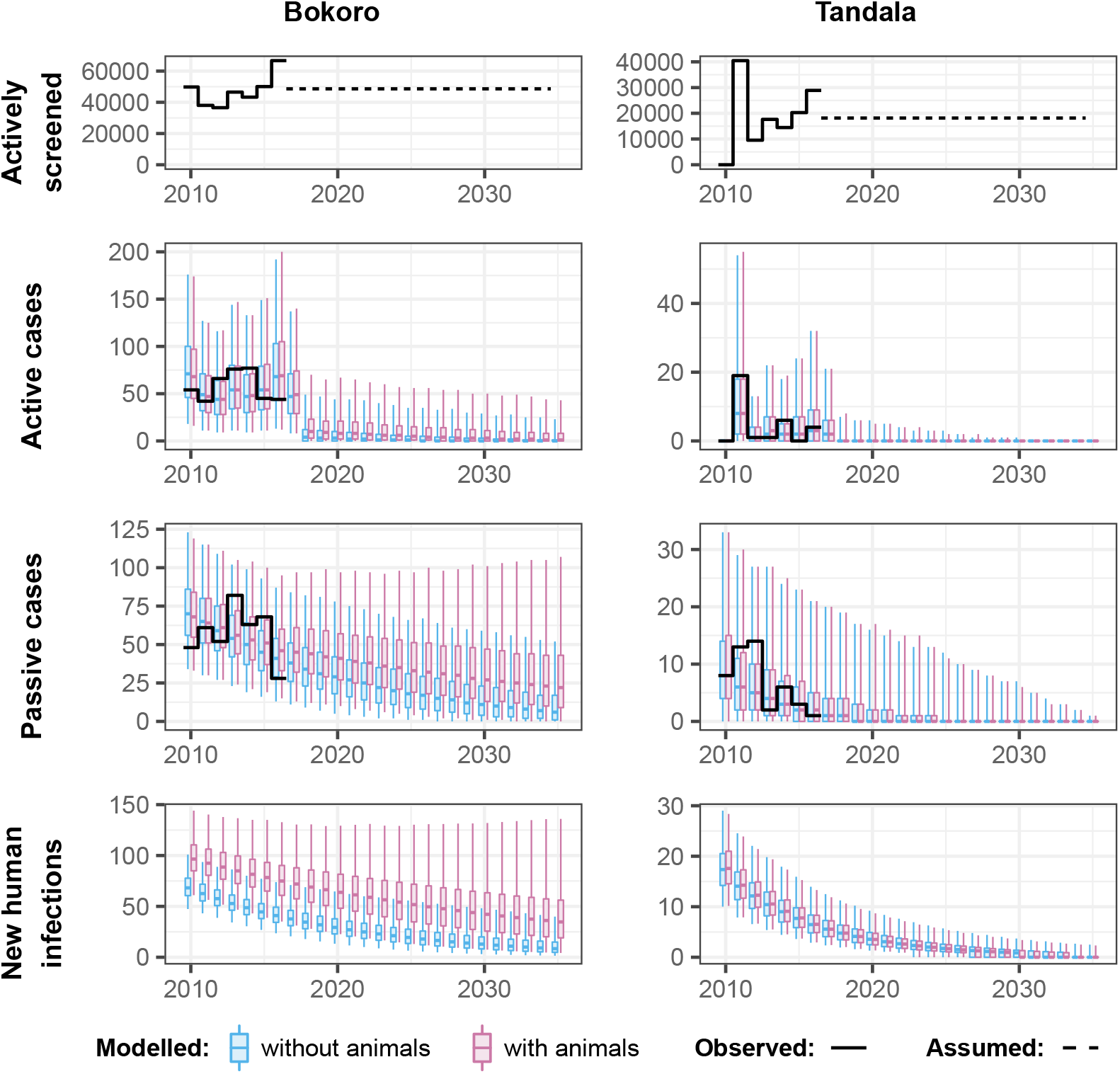
Forward projections for Bokoro and Tandala health zones for models with and without animals contributing to gHAT transmission. Within each health zone, future active screening is assumed to be at the average level of screenings from 2012–2016, while passive screening continues at the 2016 level of effectiveness. In Bokoro, and other health zones in the former province of Bandundu, the specificity of active screening was assumed to increase to 100% in 2018.

Our model results suggest that Bokoro will not reach EoT by 2030 under either model under the MeanAS strategy (see Fig 6), and if animal transmission is occurring, EoT is predicted not to be achieved for many years. However, the use of an intervention that targets transmission to and from both human and animals hosts should improve the situation. Fig 7 shows the impact that introducing vector control in Bokoro from 2020 could have on the achievement of EoT in this health zone.

**Fig 7.**
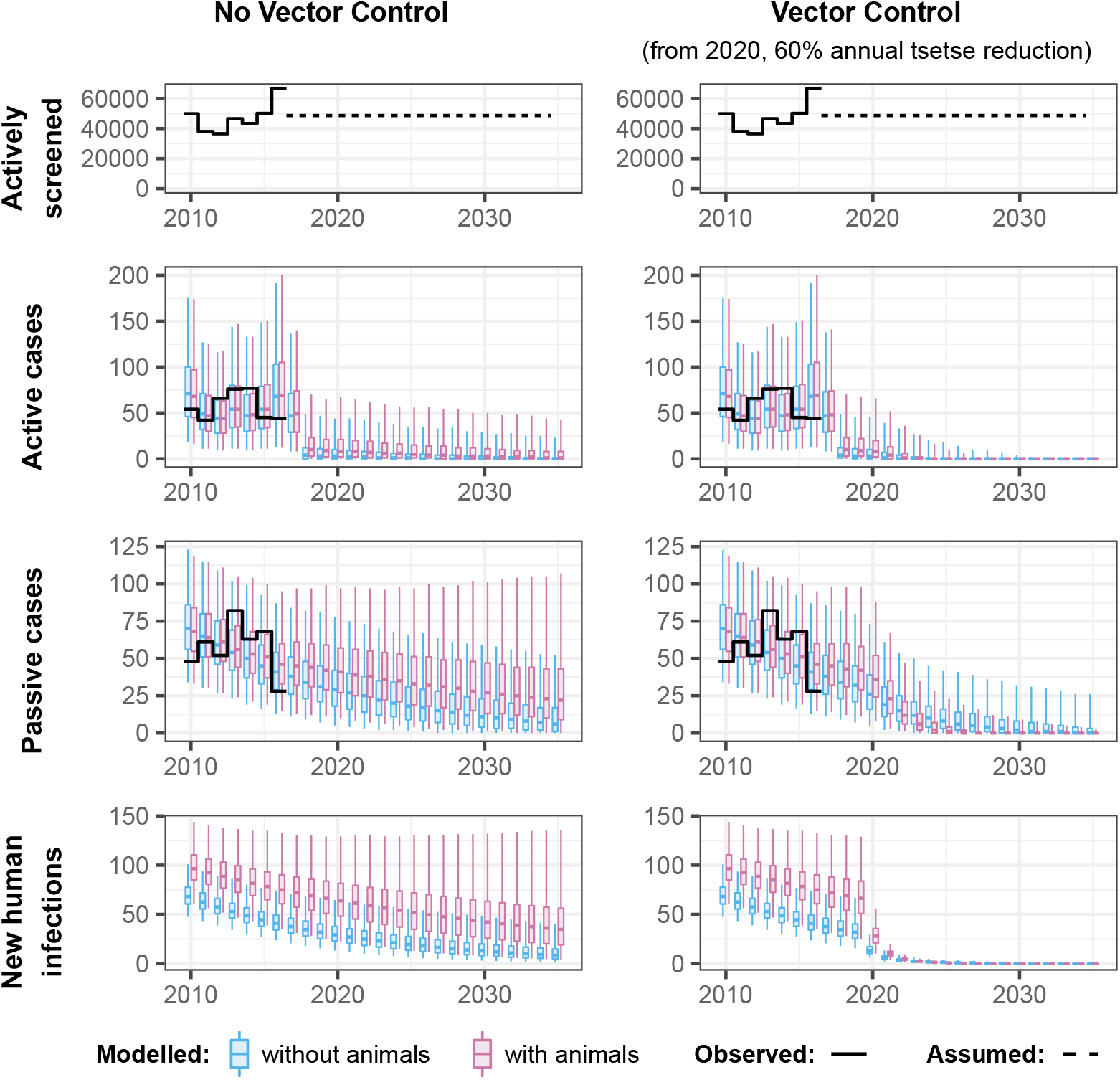
Forward projections in Bokoro health zone for models with and without an animals contributing to gHAT transmission and with or without Tiny Target-based vector control from 2020. Future active screening is assumed to be at the average level of screenings from 2012–2016. In Bokoro, and other health zones in the former province of Bandundu, the specificity of active screening was assumed to increase to 100% in 2018.

The expected number of health zones that has achieved EoT in any year was obtained by summing each of the probabilities of having having achieved EoT by that year. This was also considered for the case where VC was only carried out in health zones with less than 90% probability of having achieved EoT by 2030 without VC. These values are plotted as percentages of the 158 health zones analysed for both models and for the strategy with and without VC against year in Fig 8. An extended version of this figure is provided in the SI (S1 Materials and Methods, Fig S1.4) including the results from fitting an ensemble of the two models weighted by the model evidence. The 90% probability cut-off removes 38 (under Model 4), 33 (under Model 7) and 36 (under the ensemble model) health zones from the group in which VC was carried out. As these health zones have a high probability of reaching EoT without VC, not performing VC in these locations has only a small impact on overall progress to EoT, particularly by 2030. It is clearly desirable, in terms of resources and costs to avoid such additional interventions where they are likely unnecessary.

**Fig 8.**
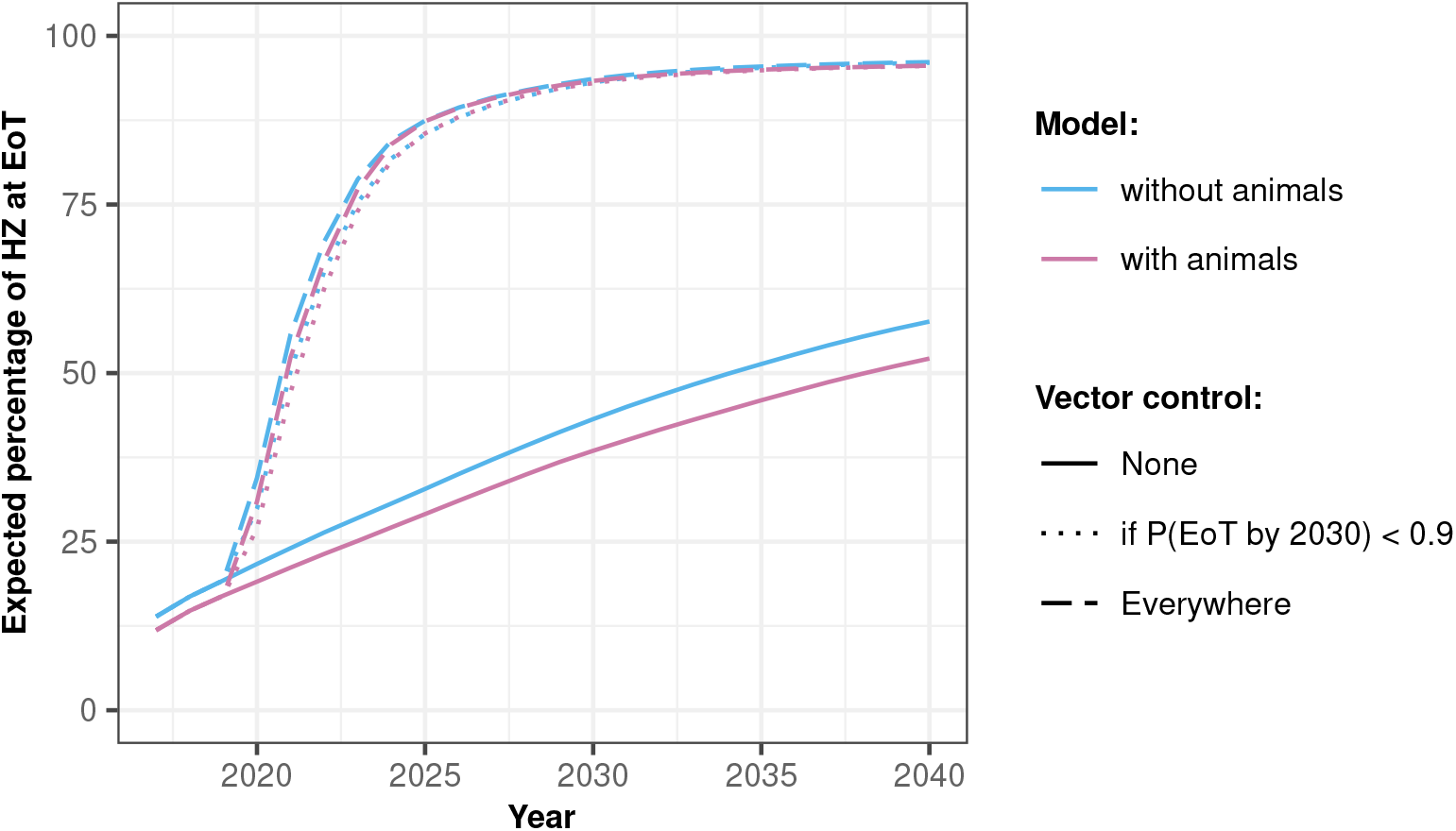
The percentage of health zones studied expected to have reached elimination of transmission (EoT) against year. Vector control (VC) was either simulated in none (solid lines), all (dashed lines), or a subset of health zones in which the probability of reaching EoT by 2030 without VC was less than 0.9 (dotted lines; using this cut-off measure, VC was simulated in 76% of health zones in the model without animal transmission and in 79% of health zones in the model with animal transmission).

Results from both models and a model evidence-based ensemble of the two are available online (https://hatmepp.warwick.ac.uk/animalfitting/v1/) for all 158 health zones analysed, including projection results for all strategies and posterior distributions of fitted parameters.

## Discussion

There are 24 health zones out of 158 in which there are substantial to decisive levels of statistical support for the model including animal transmission. In many of these health zones we identified a similar trend in the data that the health zones with support for the model with animal transmission are among those with a relatively long time series of low-level passive case detection while also having been subject to reasonable active screening coverage, if only historically. As case numbers fall, it is possible that if we were to repeat this study in a decade’s time, there would be more support for the presence of an animal reservoir as the observed data timeseries grows.

More optimistically, this study indicates that it is extremely unlikely that a maintenance animal reservoir exists in DRC – the highest estimated probability of this was 0.045, and only nine health zones had non-zero probabilities of this. In most health zones, if animals do contribute to transmission, the most probable scenario is that transmission in both humans and animals is required for maintenance of gHAT. Previous modelling in Cameroon, incorporating point estimates of animal prevalence, suggested that a combination of wild and domestic animals could constitute a reservoir [13]. The importance of our finding would be that interventions targeting transmission in humans will continue to reduce the prevalence of gHAT in DRC and EoT should be reachable, but the existence of animal transmission may delay achievement of EoT. The use of vector control or other interventions that impact transmission to and from all hosts may be highly beneficial, particularly in some regions.

In this study we have considered non-specific animal transmission, which may well consist of different species, different species population sizes, and differing levels of interaction between each animal reservoir species, tsetse and humans across health zones. The purpose of this was to enable us to consider the potential importance of transmission to and from animals based only on the available human case data. In order to consider a specific animal reservoir species, considerable data collection would be required to quantify the epidemiology of gHAT within the species and transmission between the animal species, tsetse and humans. If a suite of potential animal reservoir species were considered this recording effort would increase dramatically, and even then it would be appropriate to consider a catch-all non-specific animal reservoir as well to pick up all the species that we had neglected to study. It is very difficult to imagine a scenario in which sufficient data collection across time would allow resolution of differences between one or more specific potential animal reservoir species.

Nevertheless, measures of location-specific prevalence of *T. b. gambiense* infection in animals could still provide valuable information for future modelling analysis, especially if very high prevalence or absence of infection is observed. Meanwhile tsetse blood meal analyses could also help better parameterise models, narrow our uncertainty, and identify animal species of interest for sampling. We would recommend that data from the same locations at coinciding times for both animals and humans would best enable improved model fitting, and caution that presence of non-specific *Trypanosoma*, or even *T. brucei spp*., unfortunately tells us little about transmission of the human-infective *T. b. gambiense* to and from animals. The present study could inform targeted animal sampling to collect animal data in either health zones with inconclusive evidence for or against animal transmission with the goal of improved certainty in particular where there is a large different in projected EoT timing and the choice of model alters the strategy recommendation (see Supplementary Information S1 Materials and Methods for a bivariate choropleth map of where high levels of uncertainty intersect with large difference in the probability of meeting EoT by 2030 between Model 4 and Model 7). As human prevalence is very low, there should be an expectation that large samples sizes would be needed to have moderate probability of finding animal infection where it does exist. The average within-year prevalence estimated from the model outcomes is presented in the SI (S1 Materials and Methods, Fig S1.5) for humans, animals and tsetse in Bokoro and Tandala health zones, confirming that the prevalence in animals is even lower than that found in humans.

The use of a deterministic model in this study, particularly in health zones where low prevalence has already been achieved, does raise questions around measurement of EoT and stochastic effects. A proxy threshold for our deterministic models is used here to estimate the year of EoT (less than one new infection per year to humans). From previous gHAT modelling using stochastic versions of Model 4, we know that, even at these very low prevalences, the deterministic and stochastic outputs match very well [28, 29]. Even though our underlying transmission model is deterministic, case reporting is generated by stochastic samples around fitted values and projections. The advantage of the deterministic approach is that it allows us to calculate a log-likelihood for each potential parameter set, which is far less computationally expensive than the approximations, such as in particle filtering MCMC or approximate Bayesian computation methods, required in fully stochastic model fitting. In the future there are a variety of potential options to further incorporate stochasticity – ranging from using posterior-specific proxy EoT thresholds [30], to stochastic projections from deterministic posterior parameterisation, through to fully stochastic model fits and projections. We do not expect that the use of any of these approaches would change our overall message presented here, but would be particularly valuable when answering questions such as how current data could indicate whether or not EoT has already been met, as has been done using Model 4 [31], or surrounding the issue of resurgence following scaleback of intervention activities.

The use of model evidence for model selection or comparison is regarded as the gold standard among Bayesian statisticians. The relative complexity of the models under consideration is automatically and naturally taken account of, without the use of an explicit penalty as is the case with other information criteria. An importance sampling technique was used here to approximate the model evidence using a parametric approximation to the observed joint posterior distribution of the model parameters [22]. This makes the use of Bayes factors feasible and practical across the analysis of many health zones, as the calculation of the model evidence is a very small component of the overall analysis time (i.e. relative to the MCMC and projection of multiple intervention strategies).

## Conclusion

Health zones in which there was evidence in support of the model with animal transmission are concentrated in the former provinces of Equateur and Kasai Oriental. These health zones had low levels of on-going passive detection throughout the data period while active screening activities also took place, particularly in the early 2000s. This may suggest that if similar patterns arise in other health zones – such that they reach low prevalence but not zero case reporting – support for the animal reservoir model may increase across DRC. Despite this, the present analysis suggests that animals alone are not likely to be capable of maintaining transmission, rather the presence of animal transmission could slow down progress towards the EoT goal.

Under the model without animal transmission the expected number of health zones achieving EoT by 2030 is 43%, while it is 39% with animal transmission. If it was possible to implement vector control at a large-scale across DRC from 2020, with 60% annual reduction in tsetse population, then the predicted number of health zones reaching EoT by 2030 is 94% and 93% under models without and with animal transmission, respectively. This shows the benefit of targeting the tsetse as a means of preventing transmission to and from all hosts.

## Supporting information

Supplementary Material, Methods and Results

PRIME-NTD criteria

## Data Availability

The *gambiense* human African trypanosomiasis (HAT) data were obtained from the WHO HAT Atlas and are subject to a data sharing agreement. Interested parties should apply to WHO in order to gain access to these data. Aggregated data can be viewed alongside fitting results on the paper's companion website. Code and model outputs (posterior probability distributions of fitted parameters) are available from the OSF.

https://hatmepp.warwick.ac.uk/animalfitting/v1/

https://doi.org/10.17605/osf.io/3xadf

## Supplementary Information

**S1 Materials and Methods** More detailed description of materials and methods, and additional results and figures.

**S2 Text PRIME-NTD criteria**. Addressing the PRIME-NTD criteria for good modelling practises.

## Acknowledgments

The authors thank PNLTHA for original data collection, and WHO for data access (in the framework of the WHO HAT Atlas [17, 18]). This work was supported by the Bill and Melinda Gates Foundation (www.gatesfoundation.org) through the Human African Trypanosomiasis Modelling and Economic Predictions for Policy (HAT MEPP) project [OPP1177824] (C.H., R.E.C., P.B. and K.S.R.) and through the NTD Modelling Consortium [OPP1184344] (K.S.R. and S.E.F.S.). The funders had no role in study design, data collection and analysis, decision to publish, or preparation of the manuscript.

