## Supplementary Material, Methods and Results for "Modelling to infer the role of animals in *gambiense* human African trypanosomiasis transmission and elimination in DRC"

### **Supplementary Information**:
### Modelling to infer the role of animals in *gambiense* human African trypanosomiasis transmission and elimination in DRC
### S1 Materials and Methods

Ronald E Crump[1,2◕*], Ching-I Huang[1,2◕], Simon E F Spencer[1,3], Paul E Brown[1,2], Chansy Shampa[4], Erick Mwamba Miaka[4], and Kat S Rock[1,2]

[1]Zeeman Institute for System Biology and Infectious Disease Epidemiology Research, The University of Warwick, Coventry, U.K.
[2]Mathematics Institute, The University of Warwick, Coventry, U.K.
[3]The Department of Statistics, The University of Warwick, Coventry, U.K.
[4]PNLTHA, Kinshasa, D.R.C.

December 15, 2021

◕These authors contributed equally to this work.

#### Contents

**S1.1 Data** 2

**S1.2 Model parameterisation** 5

**S1.3 Modelling passive detection and its improvement** 7

**S1.4 Modelling vector control** 9

**S1.5 Fitting** 10
S1.5.1 Likelihood . . . . . . 10
S1.5.2 Imputation of missing numbers screened information . . . . . . 10

**S1.6 Host-specific contributions to transmission** 12

**S1.7 Model comparison** 14

**S1.8 Additional results** 14
S1.8.1 Ensemble model . . . . . . 15
S1.8.2 Prevalence of infection . . . . . . 16

#### List of Figures

S1.1 Impact of tiny targets on tsetse density . . . . . . 9
S1.2 Bivariate choropleth showing support for the models with or without animals contributing to transmission and the difference in the probability ($P_d$) of achieving EoT to humans by 2030 from these two models ("High" is more than 10% difference, "Medium" is 5-10% difference, and "Low" is less than 5% difference). Shapefiles used to produce these maps are available under an ODC-ODbL licence at `https://data.humdata.org/dataset/drc-health-data`. 15

S1.3 Maps of the probability that end of transmission in humans is achieved by 2030 for the models with and without animal transmission, and an ensemble model of these. Active screening post-2016 was assumed to be at the mean level observed 2012–2016, passive screening continued at the 2016 level of effectiveness, and no vector control was performed (except in Yasa Bonga health zone). Shapefiles used to produce these maps are available under an ODC-ODbL licence at `https://data.humdata.org/dataset/drc-health-data`. . . . . . . . . . . . . . . . . 16
S1.4 The percentage of health zones studied expected to have reached elimination of transmission (EoT) against year, for the models without animal transmission, with animal transmission, and an ensemble of these two models. Active screening post-2016 was assumed to be at the mean level observed 2012–2016, and passive screening continued at the 2016 level of effectiveness. Vector control (VC) was either simulated in none (solid lines), all (dashed lines), or a subset of health zones in which the probability of reaching EoT by 2030 without VC was less than 0.9 (dotted lines; using this cut-off measure, VC was simulated in 77% of health zones in the ensemble model, 76% of health zones in the model without animal transmission, and in 79% of health zones in the model with animal transmission). . . . . . . . . . . . . . . . . . . . . . . 17
S1.5 Mean prevalence of infection within year as a percentage of the population. Human cases are considered across disease stages and population risk group. In vectors, the prevalence of all infections and infectious (salivary gland) infections is presented. . . . . . . . . . . . . . . . . 18

### S1.1 Data

This section outlines the methods used to convert available data into a format suitable for model fitting and is the same as described in Crump *et al.* [S7].

**HAT Atlas data** The HAT Atlas data for DRC were provided in a spreadsheet format. Records were annually aggregated gHAT case records, aggregated by year, surveillance type and location as defined by multiple fields. There were 117,573 rows in this file; of which 111,408 had an entry in the geolocation (longitude and latitude) fields.

Passive surveillance records with missing or zero case numbers; and active surveillance records with both missing or zero numbers screened and missing or zero case numbers were dropped from the dataset.

This left 111,454 records (105,979 with filled geolocation fields). These records were associated with 23,424 unique combinations of former province, health zone, health area, location and territory identifiers; 20,423 of which had geolocation information and 3,001 did not. We will refer to these 24,424 geographical records as gHAT locations in this document.

**DRC Shapefile** A recent shapefile for DRC was provided by UCLA (Personal communication). The shapefile contains health zones (an organisational unit with a typical population size around 100,000) across DRC; health areas (nested within health zones, these areas are typically home to around 10,000 people) for the former province of Bandundu and part of Equateur and Haut Lomami, and post–2015 province identifiers. Former province was added to these records (post–2015 provinces being nested within former province).

**Additional geographic information** The following geographical information was obtained from the Humanitarian Data Exchange [S22]:

- a health zone shapefile from the United Nations Office for the Coordination of Humanitarian Affairs (OCHA);

Table S1.1: Number of HAT Atlas records with different combinations of former province, health zone and health area recorded.

| Recorded region identifiers: | | | Number |
|---|---|---|---|
| Former province | Health zone | Health area | |
| ✓ | ✓ | ✓ | 106823 |
| ✓ | ✓ | ✓ | 391 |
| ✓ | ✓ | | 3001 |
| ✓ | | ✓ | 14 |
| ✓ | | | 1225 |

- an OCHA file of geolocations of localities; and
- a file of geolocations of health facilities from the Global Healthsite Mapping Project.

These data were used to assist in matching and locating the gHAT data, by providing alternative spellings of names and potentially geolocations for non-geolocated gHAT locations. The locality and health facility lists were concatenated, and this enlarged locality set and the OCHA health zone map were assigned geographical identifiers as per our shapefile of choice.

**Matching HAT Atlas records to DRC shapefile** Geographical identifiers associated with the HAT Atlas/gHAT location and geographical data were sanitised to assist with matching. This involved removing diacritical marks, conversion to lowercase, collapsing whitespace within identifiers to a single space, removal of leading and trailing whitespace, converting from roman to arabic numerals, removing leading m, n or g from words where they were followed by a consonant, removing leading t from words when followed by an s, and collapsing words into a single string. In addition some specific manual edits were performed during the process as they became apparent.

Matching was then applied sequentially to the gHAT locations; such that once a match had been achieved for any given gHAT location it did not act as input to subsequent steps.

1. gHAT locations with known former province (FP), health zone (HZ), health area (HA) and geolocation were located on the UCLA and OCHA shapefiles. If the FP, HZ and HA matched the values for either the UCLA or OCHA shapefiles at that point, a match was judged to have occurred and the geolocation was accepted. This matched 3,579 of the gHAT locations.
2. gHAT locations with known former province (FP), health zone (HZ) and geolocation were located on the UCLA and OCHA shapefiles. If the FP and HZ matched the values for either the UCLA or OCHA shapefiles at that point, a match was judged to have occurred and the geolocation was accepted. This matched 13,413 of the gHAT locations. *The 16,992 gHAT locations matched in these two steps accounted for 97,520 of the HAT records (87.5%).*
3. Where a gHAT location was associated with a recent active screening event (defined as an active screening record in or after 2012 with a number screened greater than 10), the geolocation was accepted. *This matched a further 454 of the gHAT locations to give a total of 100,747 gHAT records matched (90.4%).*
4. Matching to the locality information:
   (a) if health zone and location identifier match, the locality's geolocation was assigned to the gHAT location. *Matching 44 gHAT locations, giving a total of 100,811 gHAT records matched (90.5%).*
   (b) if former province and location identifier match, the locality's geolocation was assigned to the gHAT location. *This matched 331 gHAT locations, giving a total of 101,508 gHAT records matched (91.1%).*
5. Matching to shapefile by geographic identifiers only:
   (a) Former province, health zone and health area all match. *Matched 523 gHAT locations, giving a total of 102,349 gHAT records matched (91.8%).*

(b) Former province and health zone match. *Matched 3636 gHAT locations, giving a total of 110,021 gHAT records matched (98.7%).*

(c) Former province and OCHA shapefile health zone name match. *Matched 74 gHAT locations, giving a total of 110,126 gHAT records matched (98.8%).*

(d) Former province and health area match. *Matched 60 gHAT locations, giving a total of 110,195 gHAT records matched (98.9%).*

(e) Health zone name match. *Matched 10 gHAT locations, giving a total of 110,210 gHAT records matched (98.9%).*

#### S1.2 Model parameterisation

Table S1.2: **Model parameterisation (fixed parameters).** Notation, a brief description, and the values used for fixed parameters.

| Notation | Description | Value | |
|---|---|---|---|
| $N_H$ | Total human population size in 2015 | Fixed for each health zone | [S13] |
| $\mu_H$ | Natural human mortality rate | 5.4795$\times 10^{-5}$ days$^{-1}$ | [S18] |
| $B_H$ | Total human birth rate | = $\mu_H N_H$ | |
| $\sigma_H$ | Human incubation rate | 0.0833 days$^{-1}$ | [S17] |
| $\varphi_H$ | Stage 1 to 2 progression rate | 0.0019 days$^{-1}$ | [S2, S4] |
| $\omega_H$ | Recovery rate or waning-immunity rate | 0.006 days$^{-1}$ | [S12] |
| Sens | Active screening diagnostic sensitivity | 0.91 | [S3] |
| $B_V$ | Tsetse birth rate (per capita rate of depositing new pupae) | 0.0505 days$^{-1}$ | [S16] |
| $\xi_V$ | Rate of pupal development to adult flies | 0.037 days $^{-1}$ | [S16] |
| $K$ | Pupal carrying capacity | = $111.09 N_H$ | [S16] |
| $\mathbb{P}$(pupating) | Probability of a pupa surviving to emerge as an adult fly | 0.75 | [S16] |
| $\mu_V$ | Tsetse mortality rate | 0.03 days$^{-1}$ | [S17] |
| $\sigma_V$ | Tsetse incubation rate | 0.034 days$^{-1}$ | [S8, S14] |
| $\alpha$ | Tsetse bite rate | 0.333 days$^{-1}$ | [S23] |
| $p_V$ | Probability of tsetse infection per single infective bite | 0.065 | [S17] |
| $\varepsilon$ | Reduced susceptibility factor for non-teneral (previously fed) flies | 0.05 | [S15] |
| $f_H$ | Proportion of blood-meals on humans | 0.09 | [S5] |
| $\eta_H^{\text{pre}}$ | Treatment rate from stage 1, pre-1998 | 0 | Assumed |
| disp$_{\text{act}}$ | Overdispersion parameter for active detection | $4 \times 10^{-4}$ | – |
| disp$_{\text{pass}}$ | Overdispersion parameter for passive detection | $2.8 \times 10^{-5}$ | – |
| **Parameters specific to the animal reservoir (model 7)...** | | | |
| $\mu_A$ | Natural animal mortality rate | 0.0014 days$^{-1}$ | Assumed |
| $\sigma_A$ | Animal incubation rate | 0.0833 days$^{-1}$ | [S17] |

The value of $B_V$ was chosen to maintain constant population size in the absence of vector control interventions. The value of $K$ was chosen to reflect the observed bounce back rate.

Table S1.3: **Model parameterisation (fitted parameters).** Notation, brief description, and information on the prior distributions for fitted parameters.

| Notation | Description | Prior distribution* | Percentiles of prior distribution [2.5, 50 & 97.5%] | Unit |
|---|---|---|---|---|
| $R_0$ | Basic reproduction number (NGM approach) | $1+\text{Exp}(10)$ | [1.003, 1.069, 1.369] | - |
| $r$ | Relative bites taken on high-risk humans | $1+\Gamma(3.68, 1.09)$ | [2.015, 4.654, 10.028] | - |
| $k_1$ | Proportion of low-risk people | $\text{B}(16.97, 3.23)$ | [0.6564, 0.8514, 0.9609] | - |
| $\eta_H^{\text{post}\,\dagger}$ | Treatment rate from stage 1, 1998 onwards | $\Gamma(3.54, 5.32\times10^{-5})$ | [4.59, 17.1, 42.9]$\times10^{-5}$ | days$^{-1}$ |
| $\gamma_H^{\text{post}\,\dagger}$ | Combined treatment and disease-induced death rate from stage 2, 1998 onwards | $\Gamma(2.45, 0.00192)$ | [7.59, 40.7, 121]$\times10^{-4}$ | days$^{-1}$ |
| $b_{\gamma_H^{\text{pre}}}$¶ | Relative treatment/death rate from stage 2 factor, pre-1998 | $\text{B}(1, 1)$ | [0.025, 0.500, 0.975] | - |
| Spec | Active screening diagnostic specificity | $0.998+(1-0.998)\,\text{B}(7.23, 2.41)$ | [0.9989, 0.9995, 0.9999] | - |
| $u$ | Proportion of stage 2 passive cases reported | $\text{B}(20, 40)$ | [0.2208, 0.3315, 0.4564] | - |
| $d_{\text{change}}$‡ | Midpoint year for passive improvement | $2000+(2017-2000)\,\text{B}(5, 6)$ | [2003.2, 2007.7, 2012.5] | Year |
| $\eta_{H_{\text{amp}}}$§ | Relative improvement in passive stage 1 detection rate | $\Gamma(2.013, 1.049)$ | [0.258, 1.775, 5.870] | - |
| $\gamma_{H_{\text{amp}}}$§ | Relative improvement in passive stage 2 detection rate | $\Gamma(1.001, 5)$ | [0.127, 3.471, 18.455] | - |
| $d_{\text{steep}}$§ | Speed of improvement in passive detection rate | $\Gamma(39.57, 0.0270)$ | [0.761, 1.058, 1.424] | years$^{-1}$ |
| **Parameters specific to the animal reservoir (model 7)...** | | | | |
| $f_A$ | Proportion of blood meals on reservoir animals | Flat over range 0–1 | $[0.025, 0.5, 0.975]$ | - |
| $k_A$ | Relative size of animal reservoir population | $\Gamma(1.26, 19.3)$ | $[1.18, 18.3, 81.4]$ | - |

*Where Exp(.), $\Gamma(.)$ and B(.) are the exponential, gamma (parameterised with shape and scale) and beta distributions, respectively.

†Former province-specific priors used for $\eta_H^{\text{post}}$ and $\gamma_H^{\text{post}}$; prior distributions and percentiles for Bandundu presented, see Table S1.4 for other former provinces.

¶The combined treatment and disease-induced death rate from stage 2, before 1998, was: $\gamma_H^{\text{pre}} = b_{\gamma_H^{\text{pre}}}\gamma_H^{\text{post}}$.

‡$d_{\text{change}}$ is only fitted in the former province of Bandundu.

§$\eta_{H_{\text{amp}}}$, $\gamma_{H_{\text{amp}}}$ and $d_{\text{steep}}$ are only fitted in the former provinces of Bandundu and Bas Congo; the prior distributions and percentiles presented here relate to Bandundu, see Table S1.4 for Bas Congo.

#### S1.3 Modelling passive detection and its improvement

Our new model fitting (for Model 7) follows the same methods for modelling passive detection and its improvement as Crump *et al.* [S7] and this is described below for completeness.

There are two sources of passive detection improvements considered in our model: a rapid improvement due to the introduction of the card agglutination test for trypanosomes (CATT) test in all health zones in 1998 and a gradual improvement over time in the former Bandundu and Bas Congo provinces around 2008 and mid-2015 respectively. Prior distributions and percentiles of parameters related to passive detection and it's improvement over time are summarised in Table S1.4.

Improvement in passive surveillance systems over time is considered across the whole of Bandundu and Bas Congo. The province level staging data in Bandundu suggested that passive surveillance systems in Bandundu have improved over time, which was confirmed by PNLTHA and is also supported by previous modelling work [S1]. In Bas Congo, FIND implemented the use of rapid diagnostic tests (RDT) from 2015. Staging information which was available at the province–level from 2000–2012 from the paper of Lumbala *et al.* [S11], and in the HAT Atlas data for 2015 and 2016. To inform the health zone-level analyses, a province level fit was carried out to the staged case data of Lumbala *et al.* augmented with the HAT Atlas data aggregated to the former province level for the years 2013–2016. These analyses provided no evidence for improvement in passive surveillance systems (in line with the simple sigmoidal model assumed) for any provinces other than Bandundu and Bas Congo. For Bandundu and Bas Congo, gamma distributions were fitted to the province level posterior samples of $\eta_{H_{\text{amp}}}$, $\gamma_{H_{\text{amp}}}$ and $d_{\text{steep}}$. The shape $(k)$ and scale $(\theta)$ for these fitted distributions were used as the parameters of gamma prior distributions of $\eta_{H_{\text{amp}}}$, $\gamma_{H_{\text{amp}}}$ and $d_{\text{steep}}$ in all health zones of Bandundu and Bas Congo. In Bandundu health zones a scaled and shifted beta distribution was used as the prior for $d_{\text{change}}$. The use of a broader prior for $d_{\text{change}}$ than would have resulted from using the province–level posterior distribution resulted from comparing aggregate health zone-level results with province-level observed data. In Bas Congo health zones a fixed value of 2015.5 was used for $d_{\text{change}}$.

Priors for the health zone-level $\eta_H^{\text{post}}$ and $\gamma_H^{\text{post}}$ parameters were also informed by the province-level fits. Gamma prior distributions were used which had the same mode $(\text{mode} = (k-1)\,\theta)$ as Gamma distributions fitted to the province-level posterior distribution of the parameters; and a standard deviation $(s.d. = \sqrt{k}\theta)$ of $3 \times 10^{-3}$ for $\eta_H^{\text{post}}$ and $1 \times 10^{-4}$ for $\gamma_H^{\text{post}}$, being arbitrarily selected higher variation than the province-level posterior distributions.

Table S1.4: **Parameterisation of passive detection improvement.** Notation and brief description of fitted parameters related to passive detection improvement plus their within former province prior distributions and [2.5th, 50th & 97.5th] percentile.

| Parameter | | | |
|---|---|---|---|
| | **Province** | **Prior distribution** | **Percentiles of prior distribution** |
| $\eta_H^{\text{post}}$ – Treatment rate from stage 1, 1998 onwards | | | |
| | **Bandundu** | $\Gamma(3.54, 5.32 \times 10^{-5})$ | [4.59, 17.1, 42.9]$\times 10^{-5}$ |
| | **Bas Congo** | $\Gamma(12.0, 2.89 \times 10^{-5})$ | [1.78, 3.36, 5.68]$\times 10^{-4}$ |
| | **Equateur** | $\Gamma(4.92, 4.51 \times 10^{-5})$ | [7.12, 20.7, 45.7]$\times 10^{-5}$ |
| | **Kasai Occidental** | $\Gamma(10.9, 3.03 \times 10^{-5})$ | [1.64, 3.20, 5.53]$\times 10^{-4}$ |
| | **Kasai Oriental** | $\Gamma(2.90, 5.87 \times 10^{-5})$ | [3.38, 15.1, 41.5]$\times 10^{-5}$ |
| | **Katanga** | $\Gamma(1.29, 8.79 \times 10^{-5})$ | [5.88, 86.2, 376]$\times 10^{-6}$ |
| | **Kinshasa** | $\Gamma(1.26, 8.91 \times 10^{-5})$ | [5.44, 84.4, 376]$\times 10^{-6}$ |
| | **Maniema** | $\Gamma(4.25, 4.85 \times 10^{-5})$ | [5.90, 19.0, 44.3]$\times 10^{-5}$ |
| | **Orientale** | $\Gamma(1.16, 9.27 \times 10^{-5})$ | [4.24, 79.0, 373]$\times 10^{-6}$ |
| $\gamma_H^{\text{post}}$ – Treatment rate from stage 2, 1998 onwards | | | |
| | **Bandundu** | $\Gamma(2.45, 1.92 \times 10^{-3})$ | [7.59, 40.7, 121]$\times 10^{-4}$ |
| | **Bas Congo** | $\Gamma(1.48, 2.47 \times 10^{-3})$ | [2.54, 28.7, 114]$\times 10^{-4}$ |
| | **Equateur** | $\Gamma(1.95, 2.15 \times 10^{-3})$ | [4.88, 35.0, 118]$\times 10^{-4}$ |
| | **Kasai Occidental** | $\Gamma(1.71, 2.29 \times 10^{-3})$ | [3.65, 31.9, 116]$\times 10^{-4}$ |
| | **Kasai Oriental** | $\Gamma(1.49, 2.46 \times 10^{-3})$ | [2.60, 28.8, 114]$\times 10^{-4}$ |
| | **Katanga** | $\Gamma(1.53, 2.43 \times 10^{-3})$ | [2.78, 29.4, 115]$\times 10^{-4}$ |
| | **Kinshasa** | $\Gamma(1.68, 2.31 \times 10^{-3})$ | [3.50, 31.5, 116]$\times 10^{-4}$ |
| | **Maniema** | $\Gamma(2.60, 1.86 \times 10^{-3})$ | [8.45, 42.3, 122]$\times 10^{-4}$ |
| | **Orientale** | $\Gamma(2.54, 1.88 \times 10^{-3})$ | [8.11, 41.7, 122]$\times 10^{-4}$ |
| $\eta_{H_{\text{amp}}}$ – Relative improvement in passive stage 1 detection rate | | | |
| | **Bandundu** | $\Gamma(2.01, 1.05)$ | [0.258, 1.77, 5.87] |
| | **Bas Congo** | $\Gamma(5.23, 1.70)$ | [2.98, 8.33, 18.0] |
| $\gamma_{H_{\text{amp}}}$ – Relative improvement in passive stage 2 detection rate | | | |
| | **Bandundu** | $\Gamma(1.001, 5)$ | [0.127, 3.47, 18.5] |
| | **Bas Congo** | $\Gamma(1.46, 1.26)$ | [0.126, 1.45, 5.81] |
| $d_{\text{steep}}$ – Speed of improvement in passive detection rate | | | |
| | **Bandundu** | $\Gamma(39.6, 2.70 \times 10^{-2})$ | [0.761, 1.06, 1.42] |
| | **Bas Congo** | $\Gamma(3.21, 1.45)$ | [1.03, 4.18, 10.9] |

#### S1.4 Modelling vector control

In the present study we utilise the same method to simulate the impact of biannual vector control on tsetse populations as presented elsewhere [S16, S7, S10]. Fig. S1.1 shows the assumed dynamics of tsetse populations where targets are either moderately effective, with a 60% reduction (lower than in Guinea [S6]), or highly effective, with a 90% population density reduction in a year (as seen in Uganda [S19] and Yasa Bonga health zone in DRC [S20]). In this analysis we used the 90% reduction formulation in Yasa Bonga health zone as part of fitting, but 60% elsewhere for the main text results as a low, conservative estimate of what large-scale Tiny Target deployments might achieve. Results using more optimistic reductions of 80% and 90% are available online through our graphical user interface (GUI; `https://hatmepp.warwick.ac.uk/animalfitting/v1/`).

The function which describes the probability of both hitting a target and dying is time dependent (days) from when the targets where placed:

$$f_T(t) = f_{\text{max}} \left( 1 - \frac{1}{1 + \exp(-0.068(\text{mod}(t, 182.5) - 127.75))} \right) \quad \text{(S1.4.1)}$$

and $f_{\text{max}}$ is chosen such that the tsetse population after one year is at the observed/assumed percentage reduction. For the simplified model this is given by $f_{\text{max}} = 0.0305$ for a 60% reduction and $f_{\text{max}} = 0.0750$ for a 90% reduction.

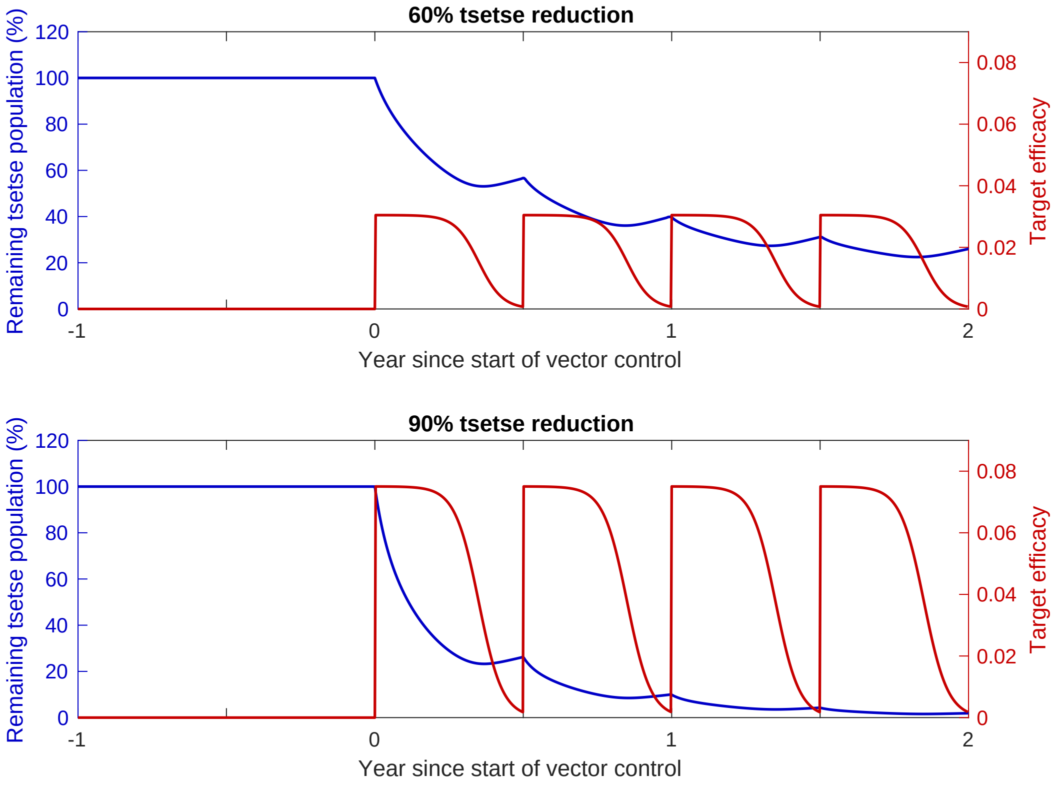

Figure S1.1: **Impact of Tiny Targets on tsetse density.** The figures show the how varying target efficacy (red line) impacts tsetse population density (blue line). Target efficacy is measured as the proportion of a host-seeking tsetse which will both hit the Tiny Target and die as a result. The graphs, reproduced from [S16], show the necessary efficacy of targets needed to reduce density by 60% (top) and 90% (bottom) by the end of the first year.

#### S1.5 Fitting

##### S1.5.1 Likelihood

Eight parameters; $R_0, r, \eta_H, \gamma_H, b_{\gamma_{H0}}, k_1, u$, and $\mathrm{Spec}$ were fitted in all health zones for both Models 4 and 7. A further two parameters; $k_A$ and $f_A$, were fitted in all health zones for Model 7. Additional parameters were included as required (combinations of $d_{\mathrm{change}}$, $\eta_{H_{\mathrm{amp}}}$, $\gamma_{H_{\mathrm{amp}}}$, $d_{\mathrm{steep}}$ and $b_{\mathrm{specificity}}$ as appropriate, see above).

The Metropolis-Hastings MCMC used a log-likelihood function:

$$\begin{aligned} LL(\theta|x) = &\ \log(P(x|\theta)) \\ \propto &\sum_{i=2000}^{2016} \left( \log \left[ \mathrm{BetaBin}\left( A_{D1}(i) + A_{D2}(i); z(i), \frac{A_{M1}(i) + A_{M2}(i)}{N_H}, \mathrm{disp}_{\mathrm{act}} \right) \right] \right. \\ &+ \log \left[ \mathrm{Bin}\left( A_{D1}(i); A_{D1}(i) + A_{D2}(i), \frac{A_{M1}(i)}{A_{M1}(i) + A_{M2}(i)} \right) \right] \\ &+ \log \left[ \mathrm{BetaBin}\left( P_{D1}(i) + P_{D2}(i); N_H, \frac{P_{M1}(i) + P_{M2}(i)}{N_H}, \mathrm{disp}_{\mathrm{pass}} \right) \right] \\ &+ \left. \log \left[ \mathrm{Bin}\left( P_{D1}(i); P_{D1}(i) + P_{D2}(i), \frac{P_{M1}(i)}{P_{M1}(i) + P_{M2}(i)} \right) \right] \right) \end{aligned}$$

The model takes parameterisation $\theta$, $x$ is the data, $P_{Dj}(i)$ and $A_{Dj}(i)$ are the number of passive/active cases (of stage $j$) in year $i$ of the data, $P_{Mj}(i)$ and $A_{Mj}(i)$ are the number of passive/active cases (of stage $j$) in year $i$ of the model, and $z(i)$ is the number of people screened in year $i$. $\mathrm{BetaBin}(m; n, p, \rho)$ gives the probability of obtaining $m$ successes out of $n$ trials with probability $p$ and overdispersion parameter $\rho$. The overdispersion accounts for larger variance than under the binomial. The probability density function of this distribution is given by:

$$\mathrm{BetaBin}(m; n, p, \rho) = \frac{\Gamma(n+1)\Gamma(m+a)\Gamma(n-m+b)\Gamma(a+b)}{\Gamma(n-m+1)\Gamma(n+a+b)\Gamma(a)\Gamma(b)}$$

where $a = p(1/\rho - 1)$ and $b = a(1-p)/p$.

##### S1.5.2 Imputation of missing numbers screened information

There are instances in the data where the number of cases from active screening within year $t$ ($A_D(t) = A_{D1} + A_{D2}$) is not consistent with the number of people recorded as having been screened in that year for that health zone ($z(t)$), i.e. $A_D(t) > z(t)$. In this situation, if $A_D(t) < 20$ we assume that these people have attended a screening outside their home health zone and the record has been correctly allocated to their home health zone and we set $z(t) = A_D(t)$. However, where $A_D(t) \geq 20$ we assume that a screening must have taken place in the health zone but that only the positive test results have been recorded. To allow use of these records we impute the missing negative test results ($A_D^-(t)$) during the MCMC analysis.

We use a hierarchical prior conditional on the other model parameters ($\theta$) for $A_D^-(t)$; $A_D^-(t)|\theta \sim \mathrm{NB}(A_D(t), P^+(\theta))$, where $P^+(\theta)$ is the probability of a positive active screening result given the current $\theta$ (obtained from solutions of the ODE up to this point in time). The proposal distribution for $A_D^-(t)$ is the same as the conditional prior, and these cancel from the posterior probability. The value $\hat{z}(t) = A_{D1} + A_{D2} + A_D^-(t)$ is used in place of the unknown $z(t)$ in the likelihood calculation.

Table S1.5: **Imputation of negative active screening results.** Former province, health zone, year and active case numbers $(A_D(t))$ where imputation of $A_D^-(t)$ was performed; plus median and 95% credible intervals of posterior distributions of $A_D^-(t)$.

| Former province | Health zone | Year $(t)$ | $A_D(t)$ | $A_D^-(t)$ | |
|---|---|---|---|---|---|
| | | | | Median | [95% CI] |
| Kinshasa | Maluku 2 | 2000 | 30 | 6052 | [4020, 9101] |
| | | 2001 | 102 | 19653 | [13792, 28992] |
| | Mont Ngafula 1 | 2000 | 303 | 214019 | [125975, 277536] |
| | Nsele | 2001 | 147 | 98902 | [70301, 149644] |
| Orientale | Doruma | 2007 | 320 | 17996 | [15550, 20466] |
| | | 2008 | 152 | 13668 | [10906, 16676] |
| | | 2011 | 215 | 40324 | [29970, 48452] |

#### S1.6 Host-specific contributions to transmission

For the model with animals contributing to transmission of gHAT (Model 7), basic reproduction number specific to animals and humans can be calculated. This process is carried out for each of 2 000 samples from the joint posterior distribution of the parameters resulting from an MCMC analysis performed as in [S7].

We construct a next generation matrix $\mathbf{K} = -\mathbf{T}\mathbf{\Sigma}^{-1}$, where $\mathbf{T}$ and $\mathbf{\Sigma}$ are $12 \times 12$ transmission and transition matrices with elements $t_{i,j}$ or $s_{i,j}$, respectively. The two dimensions of $\mathbf{T}$ and $\mathbf{\Sigma}$ map to compartments of the model thus: $E_{H1}$, $I_{1H1}$, $I_{2H1}$, $E_{H4}$, $I_{1H4}$, $I_{2H4}$, $E_A$, $I_A$, $E_{1V}$, $E_{2V}$, $E_{3V}$, and finally $I_V$ (i.e. low-risk humans who participate at random in active screening, high-risk humans who do not participate in active screening, reservoir animals and tsetse sectors), see the main text Fig 1. Equations for the non-zero elements of $T$ and $\Sigma$ are presented in Tables S1.6 and S1.7, respectively.

The overall system reproduction number, $R_0$, is the spectral radius (dominant eigenvalue) of $\mathbf{K}$ denoted by $\rho(\mathbf{K})$. Similarly to compute the human-specific or animal-specific reproduction numbers, $R_{0_\mathrm{H}}$ and $R_{0_\mathrm{A}}$, we use

$$R_{0_\mathrm{i}} = \rho((\mathbf{P}_v + \mathbf{P}_i)\mathbf{K}), \qquad i \in \{H, A\}$$

with

$$\mathbf{P}_v = \begin{pmatrix} \mathbb{0}^8 & & 0 \\ & \ddots & \vdots \\ 0 & \dots & \mathbb{1}^4 \end{pmatrix}$$

$$\mathbf{P}_H = \begin{pmatrix} \mathbb{1}^6 & & 0 \\ & \ddots & \vdots \\ 0 & \dots & \mathbb{0}^6 \end{pmatrix}$$

$$\mathbf{P}_A = \begin{pmatrix} \mathbb{0}^6 & & 0 \\ & \mathbb{1}^2 & \vdots \\ 0 & \dots & \mathbb{0}^4 \end{pmatrix}$$

If both $R_{0_\mathrm{H}} < 1$ and $R_{0_\mathrm{A}} < 1$, neither host species is classified as a maintenance host, but both are necessary for disease persistence. If $R_{0_\mathrm{H}} > 1$ and $R_{0_\mathrm{A}} < 1$, humans are both a maintenance host (can sustain disease without animals) and a reservoir (are necessary to have endemic disease).

Table S1.6: **Next generation matrix; non-zero elements of the transmission matrix, T.** Equations for $t_{i,j}$ under Model 7, the Warwick gHAT model variant with animals contributing to transmission.

| Transmission | Element, $t_{i,j}$ | Value |
|---|---|---|
| $I_V$ infects $E_{H1}$ | $t_{1,12}$ | $\lambda_{H1} = \alpha m_{\mathrm{eff}} f_H \frac{k_1}{k_1 + r k_4}$ |
| $I_V$ infects $E_{H4}$ | $t_{4,12}$ | $\lambda_{H4} = \alpha m_{\mathrm{eff}} f_H \frac{r k_4}{k_1 + r k_4}$ |
| $I_V$ infects $E_A$ | $t_{7,12}$ | $\lambda_A = \alpha m_{\mathrm{eff}} f_A$ |
| $I_{1H1}$ infects $E_{1V}$ | $t_{9,2}$ | $\alpha f_H \frac{k_1}{k_1 + r k_4} p_V \frac{(S_V^* + \varepsilon G_V^*)}{k_1 N_H}$ |
| $I_{2H1}$ infects $E_{1V}$ | $t_{9,3}$ | $= t_{9,2}$ |
| $I_{1H4}$ infects $E_{1V}$ | $t_{9,5}$ | $\alpha f_H \frac{r k_4}{k_1 + r k_4} p_V \frac{(S_V^* + \varepsilon G_V^*)}{k_4 N_H}$ |
| $I_{2H4}$ infects $E_{1V}$ | $t_{9,6}$ | $= t_{9,5}$ |
| $I_A$ infects $E_{1V}$ | $t_{9,8}$ | $\alpha f_A p_V \frac{(S_V^* + \varepsilon G_V^*)}{k_A N_H}$ |

See main text Fig 1 for model compartment information, Tables S1.2 and S1.3 for definitions of most parameters, and:
$m_{\mathrm{eff}}$ is the tsetse-to-human relative density (from $R_0$);
$S_V^* = \frac{\mu_V N_H}{\alpha + \mu_V}$ is the equilibrium value of $S_V$; and
$G_V^* = \frac{\alpha N_H}{\alpha + \mu_V}$ is the equilibrium value of $G_V$.

Table S1.7: **Next generation matrix; non-zero elements of the transition matrix, $\mathbf{\Sigma}$.** Equations for $s_{i,j}$ under Model 7, the Warwick gHAT model variant with animals contributing to transmission.

| Transition | Element, $s_{i,j}$ | Value |
|---|---|---|
| Leaves $E_{H1}$ | $s_{1,1}$ | $-\sigma_H - \mu_H$ |
| Enters $I_{1H1}$ from $E_{H1}$ | $s_{2,1}$ | $\sigma_H$ |
| Leaves $I_{1H1}$ | $s_{2,2}$ | $-\eta_H^{\text{pre}} - \phi_H - \mu_H$ |
| Enters $I_{2H1}$ from $I_{1H1}$ | $s_{3,2}$ | $\phi_H$ |
| Leaves $I_{2H1}$ | $s_{3,3}$ | $-\gamma_H^{\text{pre}} - \mu_H$ |
| Leaves $E_{H4}$ | $s_{4,4}$ | $-\sigma_H - \mu_H$ |
| Enters $I_{1H4}$ from $E_{H4}$ | $s_{5,4}$ | $\sigma_H$ |
| Leaves $I_{1H4}$ | $s_{5,5}$ | $-\eta_H^{\text{pre}} - \phi_H - \mu_H$ |
| Enters $I_{2H4}$ from $I_{1H4}$ | $s_{6,5}$ | $\phi_H$ |
| Leaves $I_{2H4}$ | $s_{6,6}$ | $-\gamma_H^{\text{pre}} - \mu_H$ |
| Leaves $E_A$ | $s_{7,7}$ | $-\sigma_A - \mu_A$ |
| Enters $I_A$ from $E_A$ | $s_{8,7}$ | $\sigma_A$ |
| Leaves $I_A$ | $s_{8,8}$ | $-\mu_A$ |
| Leaves $E_{1V}$ | $s_{9,9}$ | $-3\sigma_V - \mu_V$ |
| Enters $E_{2V}$ from $E_{1V}$ | $s_{10,9}$ | $3\sigma_V$ |
| Leaves $E_{2V}$ | $s_{10,10}$ | $-3\sigma_V - \mu_V$ |
| Enters $E_{3V}$ from $E_{2V}$ | $s_{11,10}$ | $3\sigma_V$ |
| Leaves $E_{3V}$ | $s_{11,11}$ | $-3\sigma_V - \mu_V$ |
| Enters $I_V$ from $E_{3V}$ | $s_{12,11}$ | $3\sigma_V$ |
| Leaves $I_V$ | $s_{12,12}$ | $-\mu_V$ |

See main text Fig 1 for model compartment information, Tables S1.2 and S1.3 for model parameter definitions.

#### S1.7 Model comparison

We compare our models on the basis of Bayes factors; $K_{ij} = \pi(\mathbf{x}|i) / \pi(\mathbf{x}|j)$, where $\pi(\mathbf{x}|m)$ is the marginal likelihood, or *evidence*, of the data $\mathbf{x}$ for model $m$. Here, $m \in \{7, 4\}$ indicating the Warwick gHAT model variants with and without animals contributing to transmission (Models 7 and 4 respectively for consistency with usage in previous publications). The use of Bayesian model evidence makes use of the full probability distribution of the model rather than a point estimate (usually the maximum likelihood estimate) and naturally accounts for differences in the number of parameters required for different models.

An importance sampled estimator of the model evidence was implemented following Touloupou *et al.* [S21].

The joint distribution of $(\boldsymbol{\theta}_m, \mathbf{x})$, for parameters $\boldsymbol{\theta}_m = (\theta_1, \theta_2, \ldots, \theta_{d_m})$ of model $m$ and data $\mathbf{x} = (x_1, x_2, \ldots, x_n)$ satisfies

$$\pi(\boldsymbol{\theta}_m|\mathbf{x})\, \pi(\mathbf{x}|\mathbf{m}) = \pi(\mathbf{x}|\boldsymbol{\theta}_m)\, \pi(\boldsymbol{\theta}_m) \tag{S1.7.1}$$

Where $\pi(\boldsymbol{\theta}_m|\mathbf{x})$ is the joint posterior distribution of parameters $1 \ldots d$; $\pi(\mathbf{x}|\mathbf{m})$ is the marginal likelihood or *evidence*; $\pi(\mathbf{x}|\boldsymbol{\theta}_m)$ is the likelihood, and $\pi(\boldsymbol{\theta}_m)$ is the prior distribution.

By use of Markov chain Monte Carlo (MCMC) methods to investigate the posterior distribution of the parameters, calculation of $\pi(\mathbf{x})$ is avoided. Calculation of the evidence for use in model comparison requires computing the integral:

$$\pi(\mathbf{x}|\mathbf{m}) = \int \pi(\mathbf{x}|\boldsymbol{\theta}_m)\, \pi(\boldsymbol{\theta}_m)\, d\boldsymbol{\theta}_m \tag{S1.7.2}$$

$$= \int \pi(\mathbf{x}|\boldsymbol{\theta}_m)\, \frac{\pi(\boldsymbol{\theta}_m)}{q(\boldsymbol{\theta}_m)} q(\boldsymbol{\theta}_m)\, d\boldsymbol{\theta}_m \tag{S1.7.3}$$

Equation S1.7.2 cannot be calculated analytically except for some small set of tractable models. It can, however, be rewritten as equation S1.7.3, where $q(\boldsymbol{\theta}_m)$ is a $d_m$-dimensional probability density function. From this, an importance sampled estimator of $\pi(\mathbf{x}|\mathbf{m})$ is:

$$\hat{P}_q = \frac{1}{N} \sum_{i=1}^{N} \pi(\mathbf{x}|\boldsymbol{\theta}_{m,i})\, \frac{\pi(\boldsymbol{\theta}_{m,i})}{q(\boldsymbol{\theta}_{m,i})} \tag{S1.7.4}$$

A defence mixture [S9] was used for $q(\boldsymbol{\theta}_m)$:

$$q(\boldsymbol{\theta}_m) = p\phi(\boldsymbol{\theta}_m; n, \boldsymbol{\mu}_1 \ldots \boldsymbol{\mu}_n, \mathbf{C}_1 \ldots \mathbf{C}_n) + (1-p)\, \pi(\boldsymbol{\theta}_m) \tag{S1.7.5}$$

Where $\phi(\cdot)$ is a mixture of $n$ multivariate Gaussian distributions with means $\boldsymbol{\mu}_j$ ($j = \{1 \ldots n\}$), and covariance matrices $\mathbf{C}_j$, and $p$ is a mixing proportion ($p = 0.95$ was chosen for use, being a typical value [S21]).

In each of our health zone-level MCMC analyses of Models 7 and 4, 2 000 samples from the joint posterior distribution were generated and stored, and $\phi(\boldsymbol{\theta}_m; n, \boldsymbol{\mu}_1 \ldots \boldsymbol{\mu}_n, \mathbf{C}_1 \ldots \mathbf{C}_n)$ for each health zone and model was chosen using the Matlab `fitgmdist` function, selecting $n$ based on Akaike's Information Criterion (AIC). To account for the high correlations between some of our model parameters, regularisation was applied to ensure that the covariance matrices, $\mathbf{C}_k$, would be positive semi-definite. Before passing to `fitgmdist`, transformations were applied to the posterior samples to put them in the range $[-\infty, \infty]$ – appropriate for Gaussian distributions – followed by scaling and centring to keep the regularisation consistent across analyses, at least at the simple, single overall covariance matrix level.

Having defined $\phi(\cdot)$ for a given analysis (health zone, model combination), $\hat{P}_q$ was calculated (equation S1.7.4) using $N = 2\,000$ samples drawn from $q(\boldsymbol{\theta}_m)$.

#### S1.8 Additional results

Fig S1.2 is a bivariate choropleth map which combines support for models with and without animal transmission with difference in the probability of achieving end of transmission by 2030 under the strategy where active screening in the health zone continues at the mean level observed in the period 2012-2016 ($P_d$ = $\mathbb{P}$(EOT by 2030|Model 4)−$\mathbb{P}$(EOT by 2030|Model 7). For presentation in Fig S1.2 both variables were put into

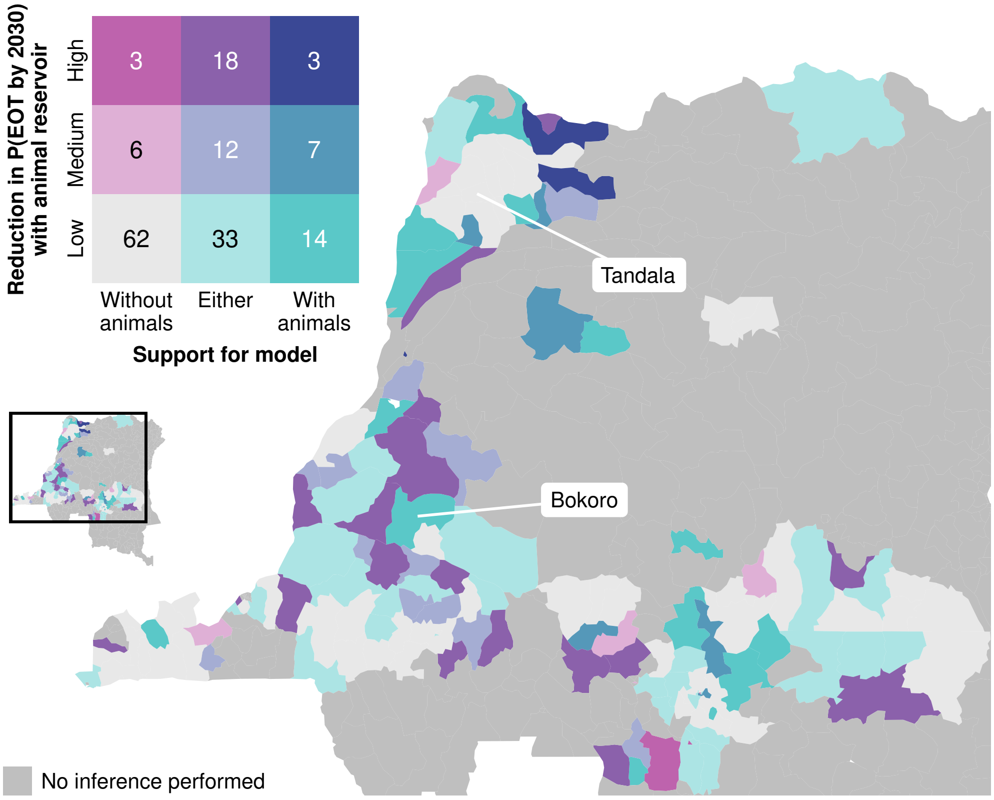

Figure S1.2: Bivariate choropleth showing support for the models with or without animals contributing to transmission and the difference in the probability ($P_d$) of achieving EoT to humans by 2030 from these two models ("High" is more than 10% difference, "Medium" is 5-10% difference, and "Low" is less than 5% difference). Shapefiles used to produce these maps are available under an ODC-ODbL licence at `https://data.humdata.org/dataset/drc-health-data`.

three categories: without animals ($K_{47} > 10^{\frac{1}{2}}$), either ($K_{47} < 10^{\frac{1}{2}} \wedge K_{74} < 10^{\frac{1}{2}}$), and with animals ($K_{74} > 10^{\frac{1}{2}}$) for model support; and for difference in probability of achieving EoT by 2030 ($P_d$): low ($P_d \leq 0.05$), medium ($0.05 < P_d \leq 0.1$), and high ($P_d > 0.1$).

There are 18 health zones (dark purple) with (i) more than 10% reduction in probability of meeting the EoT goal under Model 7 (model with animal transmission) compared to Model 4 and (ii) with weak support for either model variant. In these locations there is considerable uncertainty in whether animals contribute to transmission and this could alter policy recommendations for future strategy based on model predictions. In three health zones (dark blue) there is strong evidence for animal transmission and more than 10% reduction in probability of EoT by 2030 and so these health zones (all in former Equateur province) have more pessimistic predictions than indicated in previous modelling work [S10].

##### S1.8.1 Ensemble model

Model projections were carried out taking 10 random samples around each of 1 000 random samples for the joint posterior distribution of the model parameters. The end of transmission was detected by applying a proxy threshold to the number of new human infections within year, which is an ODE output and is not sampled. As a result, there were 1 000 for each of Models 4 and 7 relating to the end of transmission. Bayesian model averaging was used to produce an ensemble model to predict the probability of EoT was created by randomly sampling $N_4$ samples from the 1 000 Model 4 samples, and $1000 - N_4$ samples from the Model 7 samples, where $N_4 \sim \text{Bin}\left(1000, \frac{K_{47}}{K_{47}+1}\right)$.

Fig S1.3 shows the probability of achieving EoT by 2030 in each health zone, for the ensemble model and the individual models with and without animal transmission. As a result of the scale covering the full range of probability from 0 to 1 to reflect the between health zone variation in probability and the between model, within health zone differences being far smaller than this, there is little visually discernible difference between

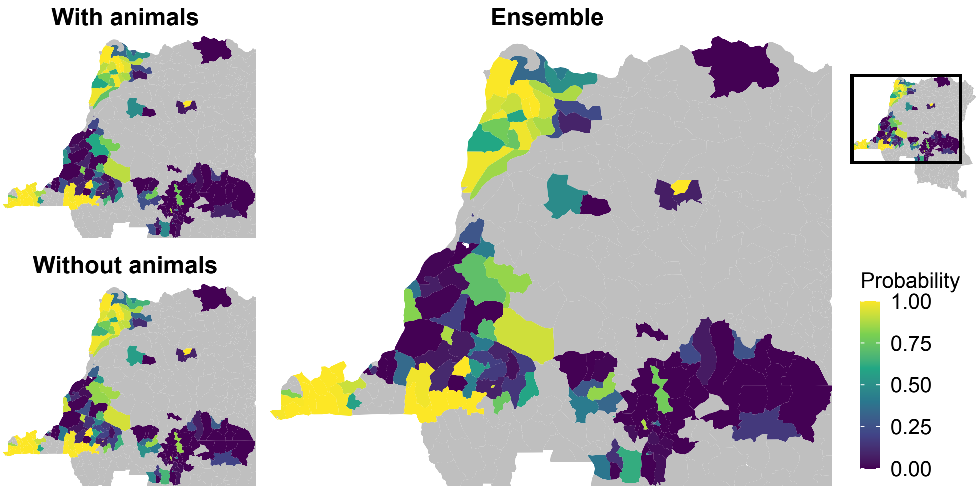

Figure S1.3: Maps of the probability that end of transmission in humans is achieved by 2030 for the models with and without animal transmission, and an ensemble model of these. Active screening post-2016 was assumed to be at the mean level observed 2012–2016, passive screening continued at the 2016 level of effectiveness, and no vector control was performed (except in Yasa Bonga health zone). Shapefiles used to produce these maps are available under an ODC-ODbL licence at `https://data.humdata.org/dataset/drc-health-data`.

the presented maps of probability, with the ensemble model being a within health zone weighted mixture of the outcomes from the other two.

The percentage of the studied health zones expected to reach EoT by the each of the years 2017–2040 is given in Fig S1.4. This is equivalent to Fig 8 in the main text, with the addition of results from the ensemble model.

##### S1.8.2 Prevalence of infection

For the model with animal transmission, the prevalence of infection was calculated for 1 000 samples from the posterior distribution of fitted parameters. The within-year mean of the number of stage 1 and 2 in low- and high-risk population groups (groups 1 and 4, respectively) ($I_{1H1}$, $I_{2H1}$, $I_{1H4}$ and $I_{2H4}$) was used for the human prevalence, and the within-year mean of $I_A$ for the animal prevalence. For vectors, we assume that infections may be detected anywhere within the tsetse (e.g. by molecular screening of the sampled tsetse), or specifically in the salivary glands (e.g. by dissection and microscopy) from where the infection can be passed on to host species. Therefore, both all infection and salivary gland infection prevalences were calculated, using the within-year mean of $E_{1V} + E_{2V} + E_{3V} + I_V$ or $I_V$.

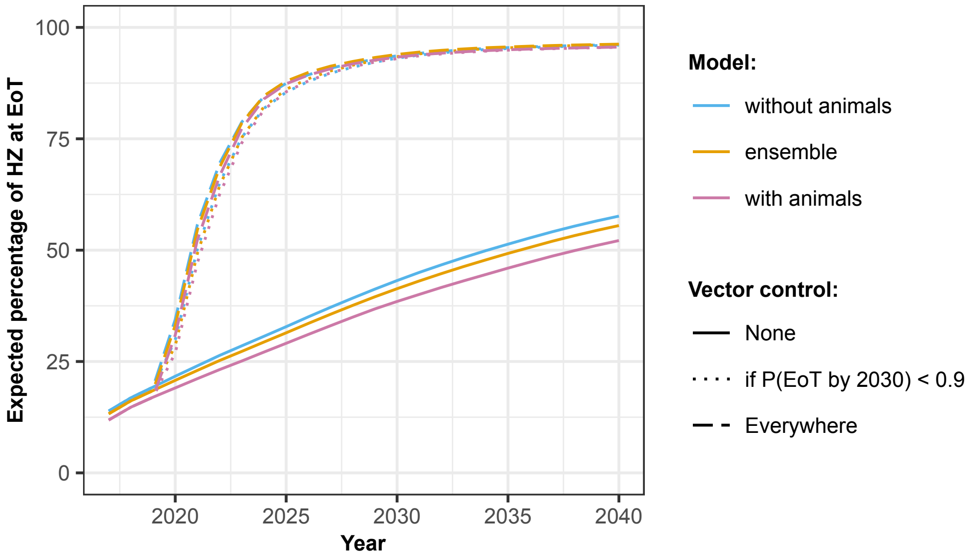

Figure S1.4: The percentage of health zones studied expected to have reached elimination of transmission (EoT) against year, for the models without animal transmission, with animal transmission, and an ensemble of these two models. Active screening post-2016 was assumed to be at the mean level observed 2012–2016, and passive screening continued at the 2016 level of effectiveness. Vector control (VC) was either simulated in none (solid lines), all (dashed lines), or a subset of health zones in which the probability of reaching EoT by 2030 without VC was less than 0.9 (dotted lines; using this cut-off measure, VC was simulated in 77% of health zones in the ensemble model, 76% of health zones in the model without animal transmission, and in 79% of health zones in the model with animal transmission).

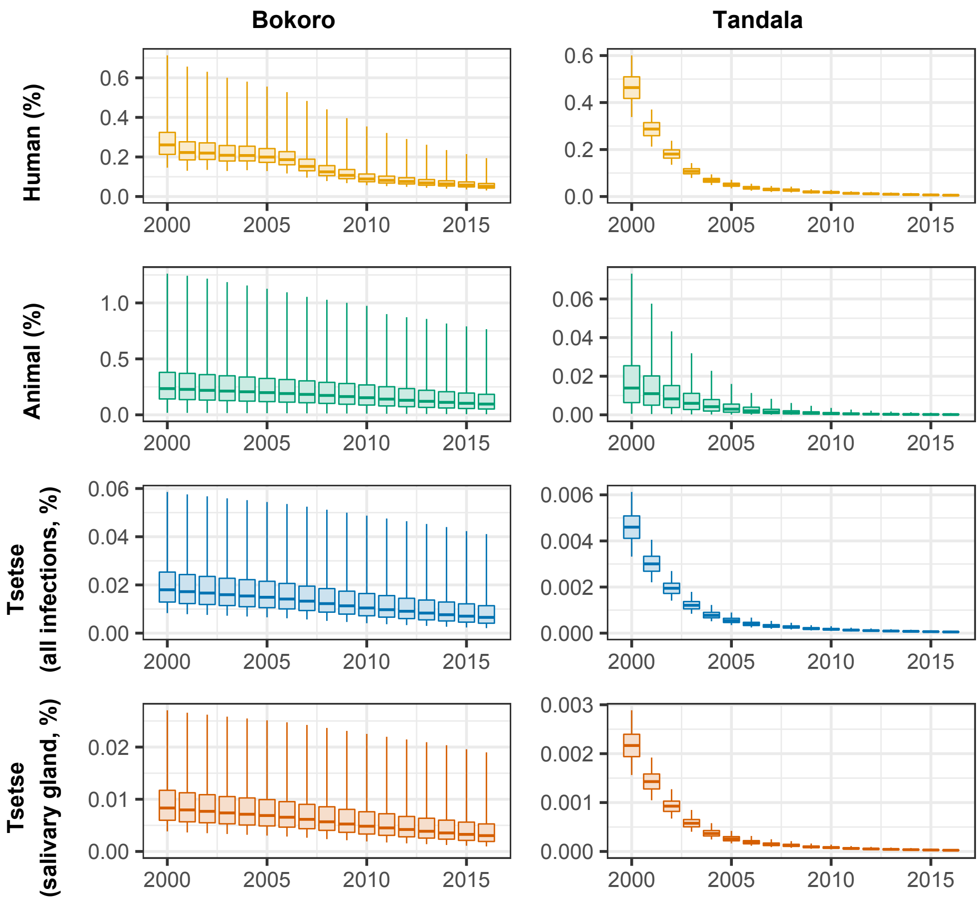

Figure S1.5: Mean prevalence of infection within year as a percentage of the population. Human cases are considered across disease stages and population risk group. In vectors, the prevalence of all infections and infectious (salivary gland) infections is presented.

[S13] OCHA Office for the Coordination of Humanitarian Affairs. *Journees Nationales de Vaccination (JNV) Activities de vaccination supplementaire, RDC*, Accessed May 2016.

[S14] S. Ravel, P. Grébaut, D. Cuisance, and G. Cuny. Monitoring the developmental status of *Trypanosoma brucei gambiense* in the tsetse fly by means of PCR analysis of anal and saliva drops. *Acta Tropica*, 88 (2):161–165, 2003.

[S15] K. S. Rock, S. J. Torr, C. Lumbala, and M. J. Keeling. Quantitative evaluation of the strategy to eliminate human African trypanosomiasis in the DRC. *Parasites & Vectors*, 8(1):532, 2015.

[S16] K. S. Rock, S. J. Torr, C. Lumbala, and M. J. Keeling. Predicting the impact of intervention strategies for sleeping sickness in two high-endemicity health zones of the Democratic Republic of Congo. *PLoS Neglected Tropical Diseases*, 11:e0005162, 2017.

[S17] D. Rogers. A general model for the African trypanosomiases. *Parasitology*, 97(1):193–212, 1988.

[S18] The World Bank. Data:Democratic Republic of Congo, 2015. URL `https://data.worldbank.org/country/congo-dem-rep?view=chart`. Accessed: 2015.

[S19] I. Tirados, J. Esterhuizen, V. Kovacic, T. N. C. Mangwiro, G. A. Vale, I. Hastings, P. Solano, M. J. Lehane, and S. J. Torr. Tsetse control and gambian sleeping sickness; implications for control strategy. *PLoS Negl Trop Dis*, 9(8):e0003822, 2015. doi: 10.1371/journal.pntd.0003822.

[S20] I. Tirados, M. C. Stanton, R. Selby, F. Mpembele, E. M. Miaka, M. Boelaert, M. J. Lehane, S. J. Torr, and A. Hope. Impact of tiny targets on glossina fuscipes quanzensis, the primary vector of human African trypanosomiasis in the Democratic Republic of the Congo. *bioRxiv*, 2020. doi: 10.1101/2020.04.07.029587. URL `https://www.biorxiv.org/content/early/2020/04/07/2020.04.07.029587`.

[S21] P. Touloupou, N. Alzahrani, P. Neal, S. E. F. Spencer, and T. J. McKinley. Efficient model comparison techniques for models requiring large scale data augmentation. *Bayesian Anal.*, 13(2):437–459, 06 2018. doi: 10.1214/17-BA1057. URL `https://doi.org/10.1214/17-BA1057`.

[S22] United Nations Office for the Coordination of Humanitarian Affairs. The humanitarian data exchange, 2018. URL `https://data.humdata.org/`. (accessed: 06.08.2018).

[S23] World Health Organization. Control and surveillance of human African trypanosomiasis: report of a WHO expert committee. Technical Report 984, World Health Organization, 2013.
