## Supplementary material for "Modelling to infer the role of animals in *gambiense* human African trypanosomiasis transmission and elimination in DRC": PRIME-NTD criteria

### **Supplementary Information**:
### Modelling to infer the role of animals in *gambiense* human African trypanosomiasis transmission and elimination in DRC
### S2 PRIME-NTD criteria

Ronald E Crump[1,2◕*], Ching-I Huang[1,2◕], Simon E F Spencer[1,3], Paul E Brown[1,2], Chansy Shampa[4], Erick Mwamba Miaka[4], and Kat S Rock[1,2]

[1]Zeeman Institute for System Biology and Infectious Disease Epidemiology Research, The University of Warwick, Coventry, U.K.
[2]Mathematics Institute, The University of Warwick, Coventry, U.K.
[3]The Department of Statistics, The University of Warwick, Coventry, U.K.
[4]PNLTHA, Kinshasa, D.R.C.

December 15, 2021

◕These authors contributed equally to this work.

#### S2.1 PRIME-NTD criteria

In order to fulfil recommendations set out for good modelling practises for policy, we itemise below how each of the five key principles relating to communication, quality and relevance of analyses - known as Policy-Relevant Items for Reporting Models in Epidemiology of Neglected Tropical Diseases (PRIME-NTD) [S1] have been met in Table S2.1.

Table S2.1: PRIME-NTD criteria fulfillment. How the NTD Modelling Consortium's "5 key principles of good modelling practice" have been met in the present study.

| Principle | What has been done to satisfy the principle? | Where in the manuscript is this described? |
|---|---|---|
| **1. Stakeholder engagement** | | |
| | This study is one in a series of modelling analyses for DRC. The modelling team lead the simulation and analysis work guided by members of the national sleeping sickness control programme in DRC (PNLTHA-DRC) – coauthors E Mwamba Miaka and S Chancy. PNLTHA-DRC have supported model development through context of how the human case data used in the study were collected and how this has changed over time (via in-person meetings, online meetings and by email). The GUI (and several variants of it) was designed in conjunction with PNLTHA-DRC to improve communication of the modelling outputs to non-modellers. It has been refined through various in-person meetings with different collaborators with the goal of providing understandable, policy-relevant outputs as well as scientific communication; over 20 non-modellers have had opportunities to interact with and provide feedback on the GUI during development. | Authorship list |
| **2. Complete model documentation** | | |
| | Full model fitting code and documentation is available through OpenScienceFramework (OSF). The model is fully described in the main text and SI. | See Materials and Methods section in the main text, Supplementary Information (file S1) and at OSF (`https://osf.io/3xadf/`) |
| **3. Complete description of data used** | | |
| | The original data and how we aggregated the data for fitting were described in detail in the SI and also in the first model fitting paper for DRC from our group [S2]. Aggregate data can be viewed next to model fits in our GUI. | See Supplementary Information (file S1, section S1.1), the GUI (`https://hatmepp.warwick.ac.uk/animalfitting/v1/`) and Crump *et al.* [S2]. |
| **4. Communicating uncertainty** | | |
| | *Structural uncertainty*: The focus of this paper is to assess statistical support for two alternative gHAT model variants – one with and one without animal transmission. In previous studies eight variants were considered [S5, S3] and there was most (and similar) support for "Model 4" and "Model 7" which are examined here. We show both fits and projections side by side to show model differences driven by this structural uncertainty<br>*Parameter uncertainty*: Parameter distributions are estimated by fitting to data. All model fits and projections include and propagate parameter uncertainty and include it in visual representations (either box and whisker plots or as probabilities, as appropriate) | *Structural uncertainty*: Materials and Methods section in main text, Figs 2, 3, 6–8.<br>*Parameter uncertainty*: All main text figures (expect Fig 1), Supplementary Information (S1) Fig S1.2 and in the GUI (`https://hatmepp.warwick.ac.uk/animalfitting/v1/`) |
| | | Continued on next page |

**Table S2.1 – continued from previous page**

| Principle | What has been done to satisfy the principle? | Where in the manuscript is this described? |
|---|---|---|
| | *Prediction uncertainty*: All predictions incorporate structural and parameter uncertainty. Furthermore, whilst we show only two main strategies in the main text, additional future interventions which are a sensitivity analysis on active screening coverage and vector control effectiveness are shown in our GUI. | *Prediction uncertainty*: GUI (`https://hatmepp.warwick.ac.uk/animalfitting/v1/`) |
| **5. Testable model outcomes** | | |
| | Previous versions of this model have undergone validation exercises (data censoring) to examine the robustness of the predictive ability of the model [S3, S4]. Whilst this was not performed here, the model is an updated version of those that have undergone validation, with updates based on critical review of model fits as data for different regions or time periods is includes (notable refinements also included here were made in the related analysis of Crump *et al.* [S2]). As more years' data become available in the future the model outputs shown here will be able to be compared to reported active and passive case data to test model predictions. | The GUI contains model predictions under various strategies for each health zone (`https://hatmepp.warwick.ac.uk/animalfitting/v1/`). |
